# Immunogenicity and safety of heterologous Omicron BA.1 and bivalent SARS-CoV-2 recombinant spike protein booster vaccines: a phase 3, randomized, clinical trial

**DOI:** 10.1101/2023.07.05.23291954

**Authors:** Chijioke Bennett, E Joy Rivers, Wayne Woo, Mark Bloch, King Cheung, Paul Griffin, Rahul Mohan, Sachin Deshmukh, Mark Arya, Oscar Cumming, A. Munro Neville, Toni McCallum Pardey, Joyce S Plested, Shane Cloney-Clark, Mingzhu Zhu, Raj Kalkeri, Nita Patel, Agi Buchanan, Alex Marcheschi, Jennifer Swan, Gale Smith, Iksung Cho, Gregory M. Glenn, Robert Walker, Raburn M. Mallory the Novavax Inc. 2019nCoV-311 Study Group

## Abstract

**Background:** Mutations present in emerging SARS-CoV-2 variants permit evasion of neutralization with prototype vaccines. A novel Omicron BA.1 subvariant-specific vaccine (NVX-CoV2515) was tested alone, or as a bivalent preparation in combination with the prototype vaccine (NVX-CoV2373), to assess antibody responses to SARS-CoV-2.

**Methods:** Participants aged 18 to 64 years immunized with 3 doses of prototype mRNA vaccines were randomized 1:1:1 to receive a single dose of NVX-CoV2515, NVX-CoV2373, or bivalent mixture in a phase 3 study investigating heterologous boosting with SARS-CoV-2 recombinant spike protein vaccines. Immunogenicity was measured 14 and 28 days after vaccination for the SARS-CoV-2 Omicron BA.1 sublineage and ancestral strain. Safety profiles of vaccines were assessed.

**Results:** Of participants who received trial vaccine (N=829), those administered NVX-CoV2515 (n=286) demonstrated superior neutralizing antibody response to BA.1 versus NVX-CoV2373 (n=274) at Day 14 (geometric mean titer ratio [95% CI]: 1.6 [1.33, 2.03]). Seroresponse rates [n/N; 95% CI] were 73.4% [91/124; 64.7, 80.9] for NVX-CoV2515 versus 50.9% [59/116; 41.4, 60.3] for NVX-CoV2373. All formulations were similarly well-tolerated.

**Conclusions:** NVX-CoV2515 elicited a superior neutralizing antibody response against the Omicron BA.1 subvariant compared with NVX-CoV2373 when administered as a fourth dose. Safety data were consistent with the established safety profile of NVX-CoV2373.

## Introduction

Since the COVID-19 outbreak in late 2019, several variants of SARS-CoV-2 (e.g., Alpha, Beta, Gamma, and Delta) have emerged with mutations in key antigenic sites in the receptor binding domain and spike protein. In late 2021, the Omicron variant emerged as the dominant circulating SARS-CoV-2 virus throughout the world, replacing earlier strains/variants. The emergence and propagation of SARS-CoV-2 variants, such as the Omicron sublineages, has complicated the COVID-19 vaccine landscape. Initial actions to stay ahead of SARS-CoV-2 evolution included directives from the US Food and Drug Administration (FDA) to develop vaccines containing an Omicron component [1,2].

Large phase 3 clinical trials for prototype COVID-19 vaccines were conducted prior to the extensive prevalence of variant strains [3-6]. High vaccine efficacy against the ancestral (Wuhan) strain of SARS-CoV-2 was reported in the phase 3 trial of BNT162b2 (Pfizer/BioNTech, July 2020-November 2020) [3], the phase 3 trial of mRNA-1273 (Moderna, July 2020-October 2020) [4], the UK-based phase 3 trial of the Matrix-M–adjuvanted, recombinant spike protein COVID-19 vaccine NVX-CoV2373 [5], and the pivotal US phase 3 trial of NVX-CoV2373 [6]. Multiple recent studies have demonstrated that newly emerging Omicron sublineages are less efficiently neutralized than the ancestral SARS-CoV-2 strain by approved prototype COVID-19 mRNA vaccines, including the BNT162b2 and mRNA-1273 vaccines [7-9]. As of June 2022, regulatory bodies have instructed COVID-19 vaccine manufacturers to develop bivalent vaccines consisting of the ancestral and Omicron BA.4/BA.5 subvariant strains to potentially provide increased protection from COVID-19 infection [1]; notably, a number of manufacturers already had bivalent vaccines consisting of the ancestral and Omicron BA.1 subvariant strains in development. Currently, the US FDA has granted emergency use authorization of two mRNA-based bivalent vaccines targeting the Omicron sublineage [10,11]; however, sparse real-world data exist about the comparative effectiveness of monovalent versus bivalent SARS-CoV-2 vaccines. In June 2023 during a Vaccines and Related Biological Products Advisory Committee (VRBPAC) Meeting, the FDA recommended development of a monovalent Omicron XBB-sublineage vaccine for the 2023–2024 vaccination campaign [12]. The World Health Organization (WHO) Technical Advisory Group on COVID- 19 Vaccine Composition (TAG-CO-VAC), as well as the European Center for Disease Prevention and Control (ECDC) and European Medicines Agency (EMA) have made similar recommendations about updating vaccines to target XBB strains [13,14].

As part of the evaluation of Omicron-targeted vaccines, Novavax produced a novel Omicron BA.1 subvariant-specific vaccine (NVX-CoV2515) based on the same recombinant spike protein technology as its authorized prototype vaccine, NVX-CoV2373. Similarly, NVX-CoV2515 is also a co-formulated product consisting of full-length, pre-fusion recombinant S protein trimers with the saponin-based adjuvant, Matrix-M™. NVX-CoV2515 was prepared for use either alone or in combination with the prototype vaccine (NVX-CoV2373) as a bivalent mixture to determine whether it would enhance or broaden antibody responses across variants of concern. To provide real-world applicable data, the population planned for investigation included participants who had already received 3 prior vaccinations with mRNA-based vaccines produced by Moderna and Pfizer BioNTech (i.e., mRNA-1273 and BNT162b2, respectively).

Here, we describe interim results from an ongoing, phase 3, randomized, observer-blinded clinical trial (NCT05372588) evaluating the immunogenicity and safety of an Omicron BA.1- containing monovalent vaccine (NVX-CoV2515) and a bivalent vaccine (NVX-CoV2373 + NVX-Cov2515) compared with the original NVX-CoV2373 booster. The goal of this interim analysis was to determine if NVX-CoV2515 induces superior antibody responses to the Omicron BA.1 subvariant compared with the antibody response induced by NVX-CoV2373.

## Methods

As part of an ongoing phase 3, randomized, observer-blinded study (NCT05372588), participants who previously received a regimen of 3 doses of mRNA-1273 and/or BNT162b2 were randomized 1:1:1 to receive NVX-CoV2373, NVX-CoV2515, or bivalent mixture as a heterologous fourth dose. Eligible participants were ≥18 and ≤64 years of age and received their last dose of mRNA vaccine ≥90 days prior to their planned study vaccination. Baseline ­­SARS-CoV-2 positivity by either polymerase chain reaction (PCR) or anti-nucleocapsid (anti-N) diagnostics was confirmed.

Participants received randomized investigational vaccines containing 5 µg SARS-CoV-2 recombinant Spike (rS) protein and 50 µg Matrix-M adjuvant administered via a 0.5 mL intramuscular injection. The bivalent vaccine was prepared on-site as a 1:1 mixture of NVX- CoV2373 and NVX-CoV2515. Following vaccination, participants utilized an electronic diary to record daily solicited local and systemic reactions for 7 days. Unsolicited adverse events (AEs) were collected for 28 days following vaccination. Data were analyzed from three participant analysis sets: the Safety Analysis Set included all participants who provided consent, were randomized, and received the study vaccine; the Per-Protocol Analysis Set 1 (PP1) population included participants who received the study vaccine, had serology results for baseline and an analyzed time point, were negative at baseline for SARS-CoV-2, and had no major protocol violations or event (e.g., COVID-19 infection) that could potentially impact immune responses; the Per-Protocol Analysis Set 2 (PP2) population was defined exactly as the PP1 population excluding the requirement for participants to have a negative baseline anti-N result. PP1 and PP2 results were determined for each strain/subvariant, serology assay, and study visit.

Immune responses were assessed at 14 and 28 days following vaccination. Participant serum collected at each timepoint was analyzed using live-virus neutralization [15], anti-spike immunoglobulin G (IgG) antibody [15], and pseudovirus neutralization assays [16]. Live-virus neutralization assays provided microneutralization with an inhibitory concentration of 50% (MN50) data for the Omicron BA.1 sublineage (Day 14, primary endpoint) and ancestral strain (validated by 360biolabs [Melbourne, Australia]). The 95% CI for geometric mean titer (GMT) and geometric mean fold rise (GMFR) were obtained based on a t-distribution of the log-transformed values. The GMT ratio (GMTR) at Day 14 and the two-sided 95% CIs were computed using the analysis of covariance (ANCOVA), with the vaccine group as the fixed effect and the titer at Day 0 as the covariate under a two-sided type I error rate of 0.05. Statistical significance was achieved if the lower-bound of the two-sided 95% CI was above unity (i.e., >1). Seroresponse rates (SRRs) in MN50 titers (defined as a ≥4-fold increase from baseline values) of the Omicron BA.1 subvariant at Day 14 following study vaccination were analyzed as part of the primary endpoint. Two-sided exact binomial 95% CIs were calculated using the Clopper-Pearson method. The difference in SRR between groups (expressed as NVX-CoV2515 minus NVX- CoV2373) was also calculated, with the 95% CI for the difference based on the method of Miettinen and Nurminen. For the analysis of difference of SRRs, assuming 80% SRR for NVX- CoV2373 and 85% SRR for NVX-CoV2515, there was 90%power to conclude non-inferiority using a margin of -5% (NVX-CoV2515 relative to NVX-CoV2373). Serum anti-spike IgG enzyme linked immunosorbent assays (ELISAs) measured participant serum antibody concentrations (EU/mL) against the recombinant spike protein of the ancestral strain and Omicron BA.1 sublineage (validated by Novavax Clinical Immunology [Gaithersburg, Maryland, USA]). As an exploratory endpoint, pseudovirus neutralization assays provided 50% inhibitory dilution (ID50) data for the ancestral strain and Omicron BA.1 sublineage (validated by Monogram Biosciences [South San Francisco, CA, USA]).

Sample size determination was based on the co-primary endpoints of MN50 GMT and SRR. For the GMTR analysis a standard deviation of 0.6 for log10-transformed neutralization titers based on data from previous studies, a 15% non-evaluable allowance, and an overall one-sided type I error of 2.5% was assumed. Study enrollment was halted ahead of reaching the planned population after re-examination of the sample size approximations suggested that the necessary number of participants had already been achieved to assess the primary endpoints.

The trial protocol was approved by the Alfred Hospital Ethics Committee (Melbourne, Victoria, Australia) and the Bellberry Human Research Ethics Committee (Adelaide, South Australia), and is registered on Clinicaltrials.gov (NCT05372588). This study was performed in accordance with the International Conference on Harmonization, Good Clinical Practice guidelines. All participants provided informed consent prior to study participation.

Randomization was managed by a contract research organization (CRO), and treatments were assigned using an Interactive Web Response System (IWRS). The block size was blinded information and known only by the CRO statistician. Predetermined site personnel were unblinded to enable vaccine preparation and administration without breaking the blind for other personnel or participants. Only blinded personnel could perform study related assessments or have participant contact for data collection after administration of study vaccine.

## Results

From 31 May 2022 to 17 July 2022, a total of 835 participants (from 19 study sites across Australia) were screened and 831 participants were randomized to one of three treatment groups. Of the randomized participants, 829 received vaccine: 286 received NVX-CoV2515, 274 received NVX-CoV2373, and 269 received bivalent vaccine (Safety Analysis Set) (**Supp Figure 1**).

Demographic and other baseline characteristics of the participants in the Safety Analysis Set were similar across all vaccine groups (**Table 1**). The median age was 41.0 to 42.0 years, the majority of participants in each group were female (52.2% [NVX-CoV2373, 143/274] to 56.1% [bivalent, 151/269]), and most participants were White (≥78.5% [NVX-CoV2515, 233/286; NVX-CoV2373, 215/274; bivalent, 220/269]) and of Australian ethnicity (≥86.1% [NVX- CoV2515, 252/286; NVX-CoV2373, 236/274; bivalent, 233/269]). Baseline SARS-CoV-2 exposure was substantial, with ≥50.9% (NVX-CoV2515, 149/286; NVX-CoV2373, 145/274; bivalent, 137/269) of participants testing positive by either PCR or anti-N serology. These participants were excluded from the primary endpoint analysis of the Per-Protocol Analysis Set 1 (PP1), however were included in complementary analyses of the Per-Protocol Analysis Set 2 (PP2) to provide data more representative of a “real world” population. Demographic and other baseline characteristics of participants in the PP1 and PP2 populations were similar to those in the Safety Analysis Set and were generally well balanced across the treatment groups (**Supp Table 1**).

**Table 1.**
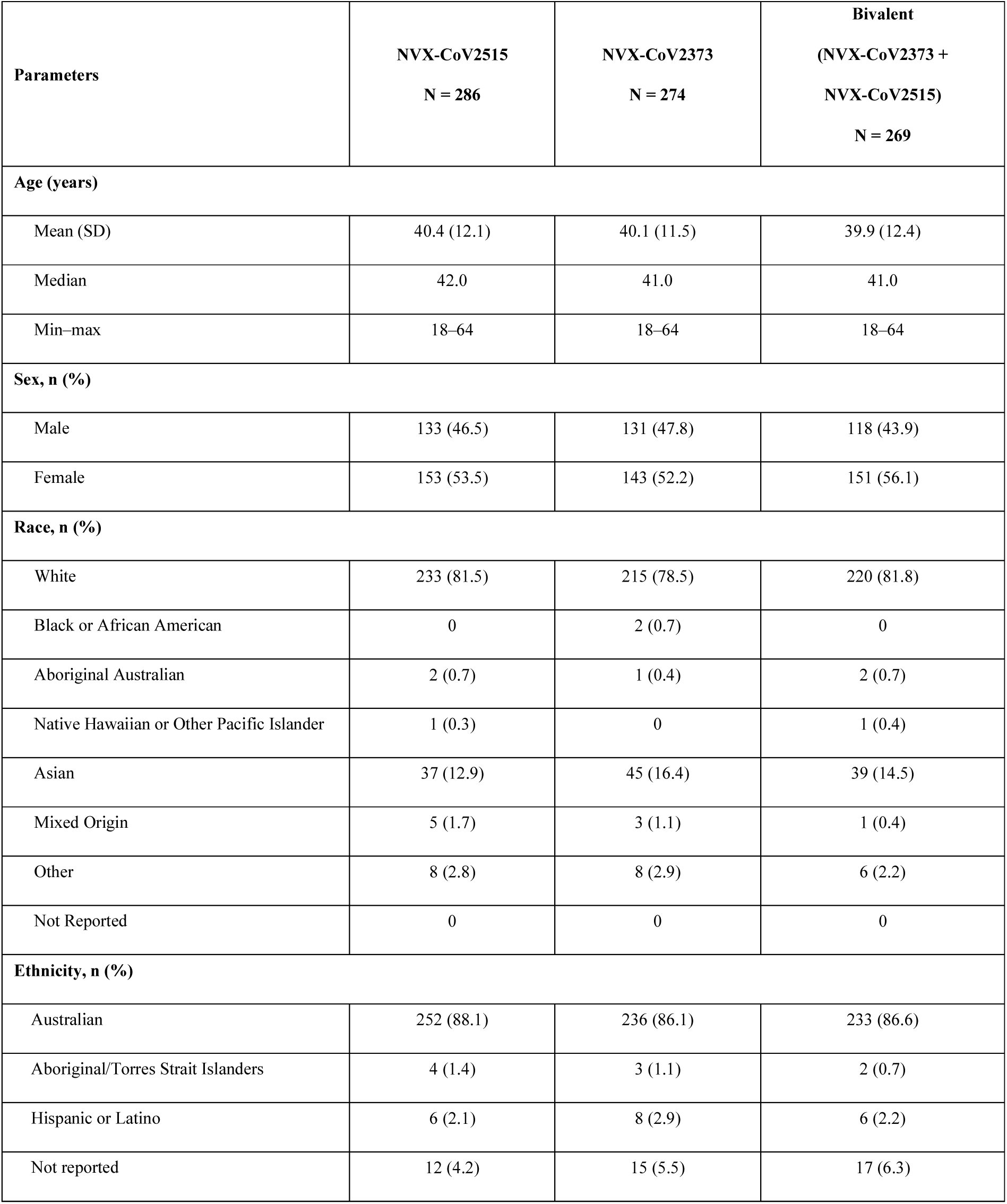
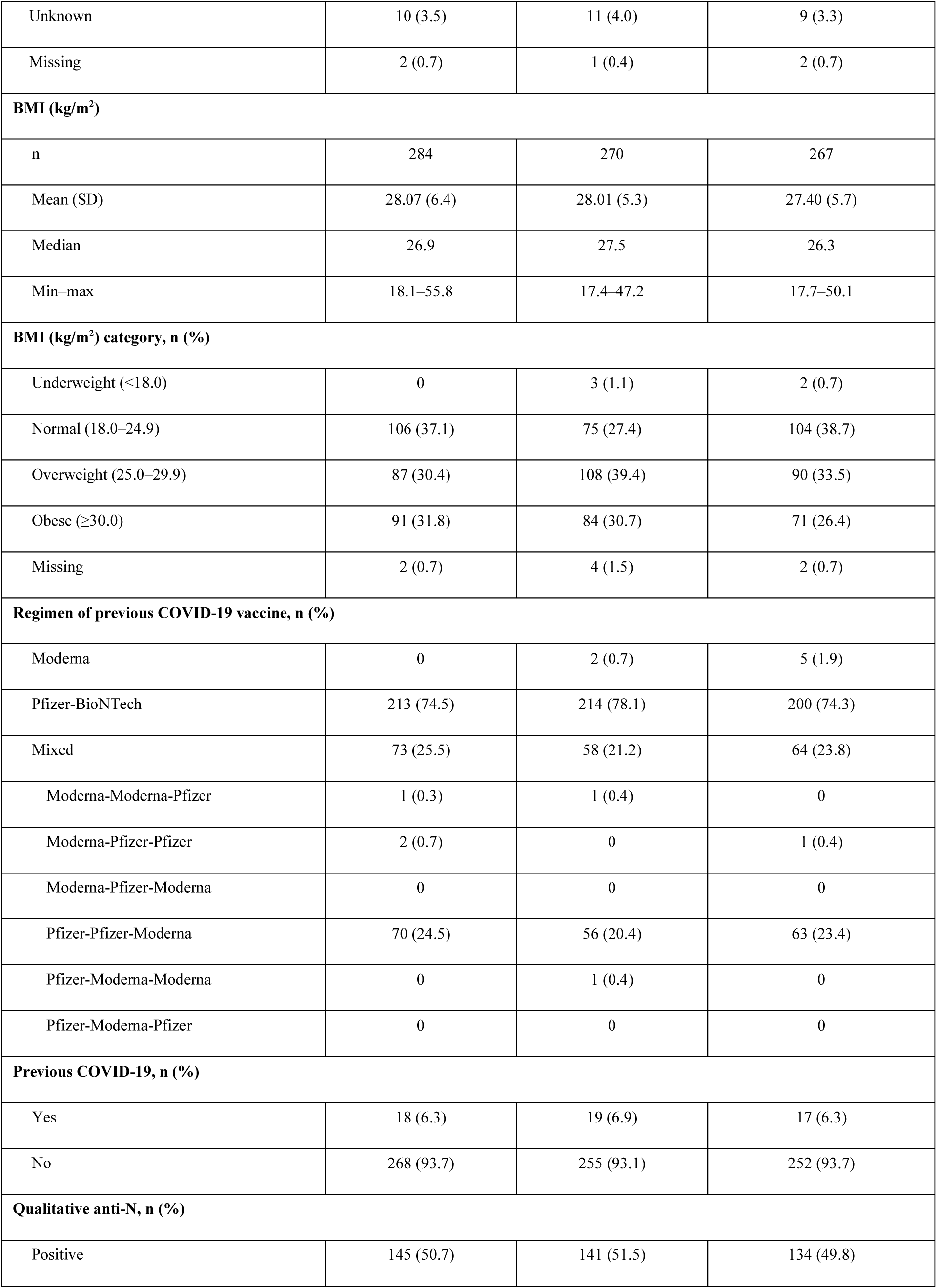
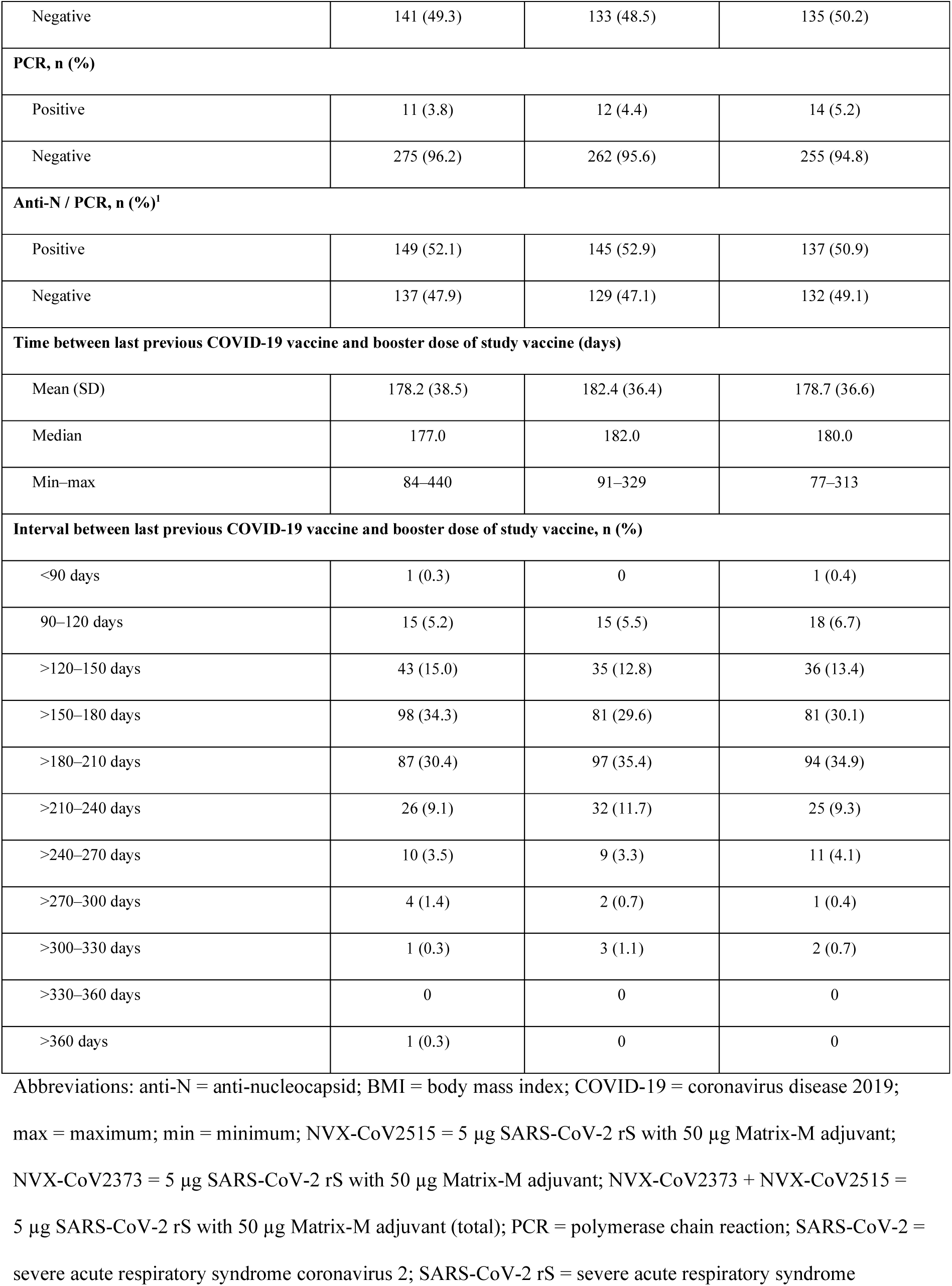
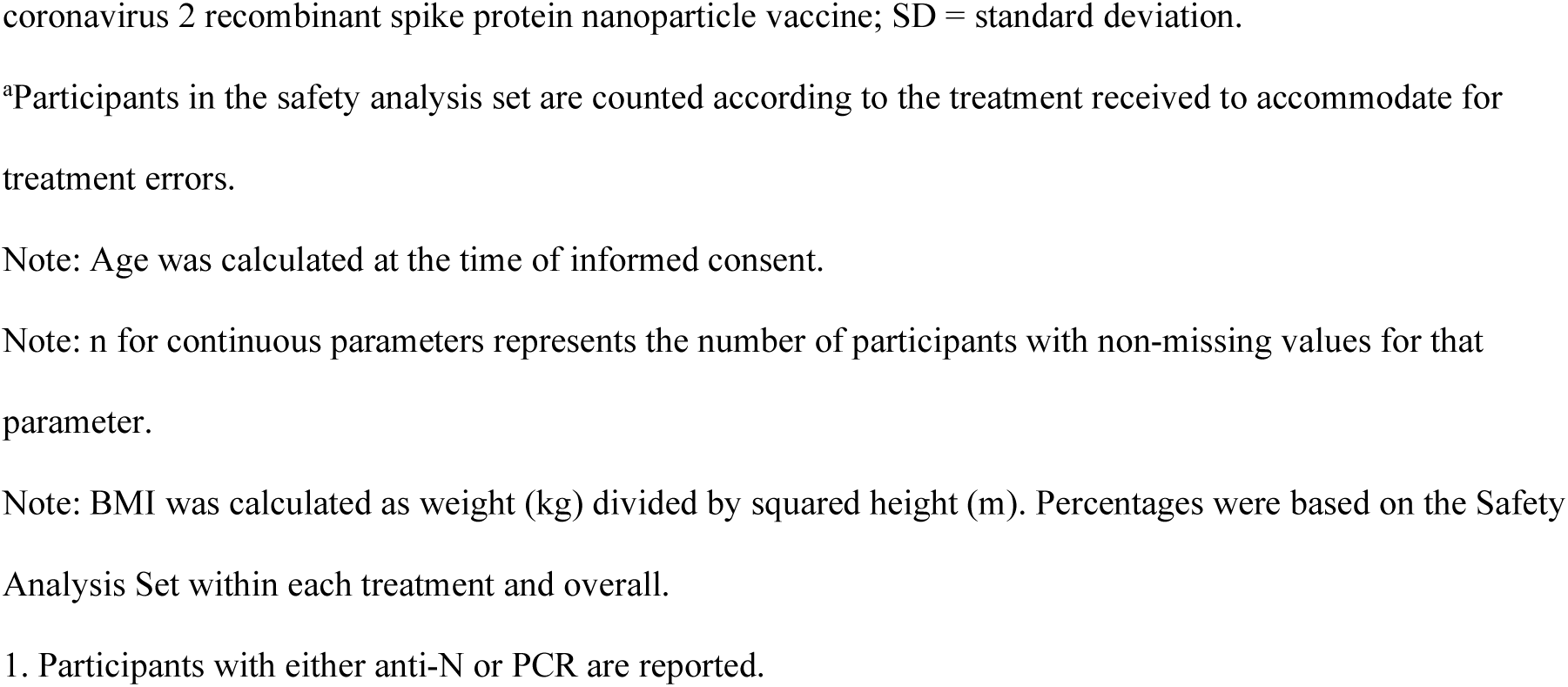
Demographics and baseline disease characteristics (Safety Analysis Set^a^)

### Per Protocol Analysis Set 1

Among participants with no evidence of SARS-CoV-2 exposure at baseline (PP1 population), strong immune responses were observed following administration with all 3 investigational vaccines at 14 days post-vaccination (**Table 2**). At Day 14, microneutralization assay GMTs against the Omicron BA.1 sublineage were the highest in participants receiving NVX-CoV2515, followed by those receiving the bivalent vaccine and NVX-CoV2373 (MN50 titers [95% CI] of 130.8 [109.2, 156.7], 97.9 [81.3, 117.9], and 83.9 [69.6, 101.2], respectively) (**Figure 1**). Formal comparison of NVX-CoV2515 and NVX-CoV2373 resulted in a GMTR of 1.6 (95% CI: 1.33, 2.03) indicating a significant difference between the vaccines (**Table 2**). NVX-CoV2515 induced a non-inferior SRR against the Omicron BA.1 subvariant virus versus NVX-CoV2373 (73.4% [91/124] vs 50.9% [59/116]) at Day 14, with a difference in SRRs of 22.5% (95% CI: 10.3, 34.2) **(Table 2)**. MN50 GMTs for the ancestral strain microneutralization assay were somewhat lower in participants receiving NVX-CoV2515 compared with the bivalent vaccine and NVX- CoV2373 alone (MN50 titers [95% CI] of 1076.3 [908.4, 1275.4], 1319.9 [1120.1, 1555.3], and 1442.5 [1192.4, 1745.0], respectively) (**Figure 1**). Similar findings were seen at Day 28.

**Figure 1.**
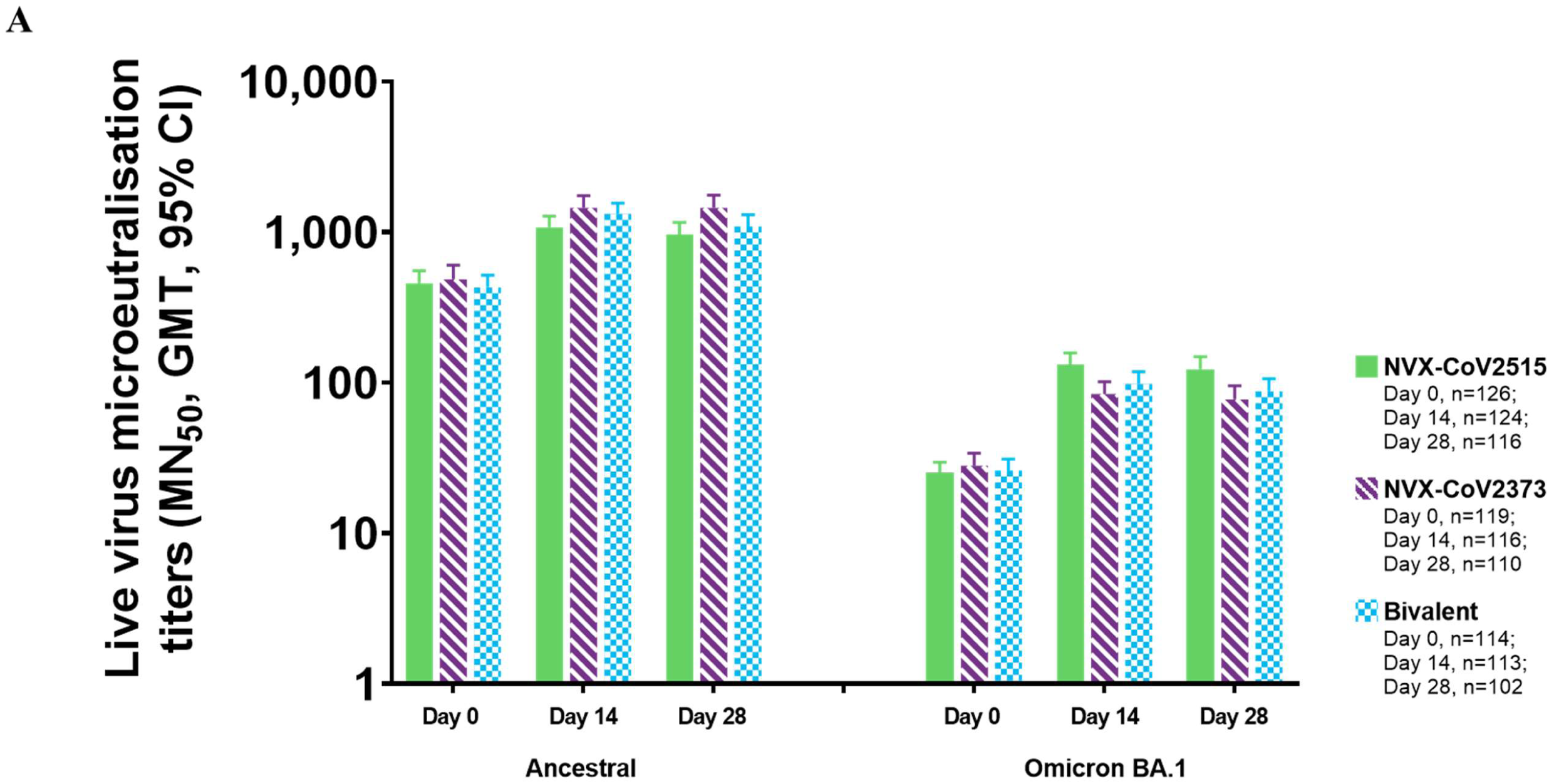
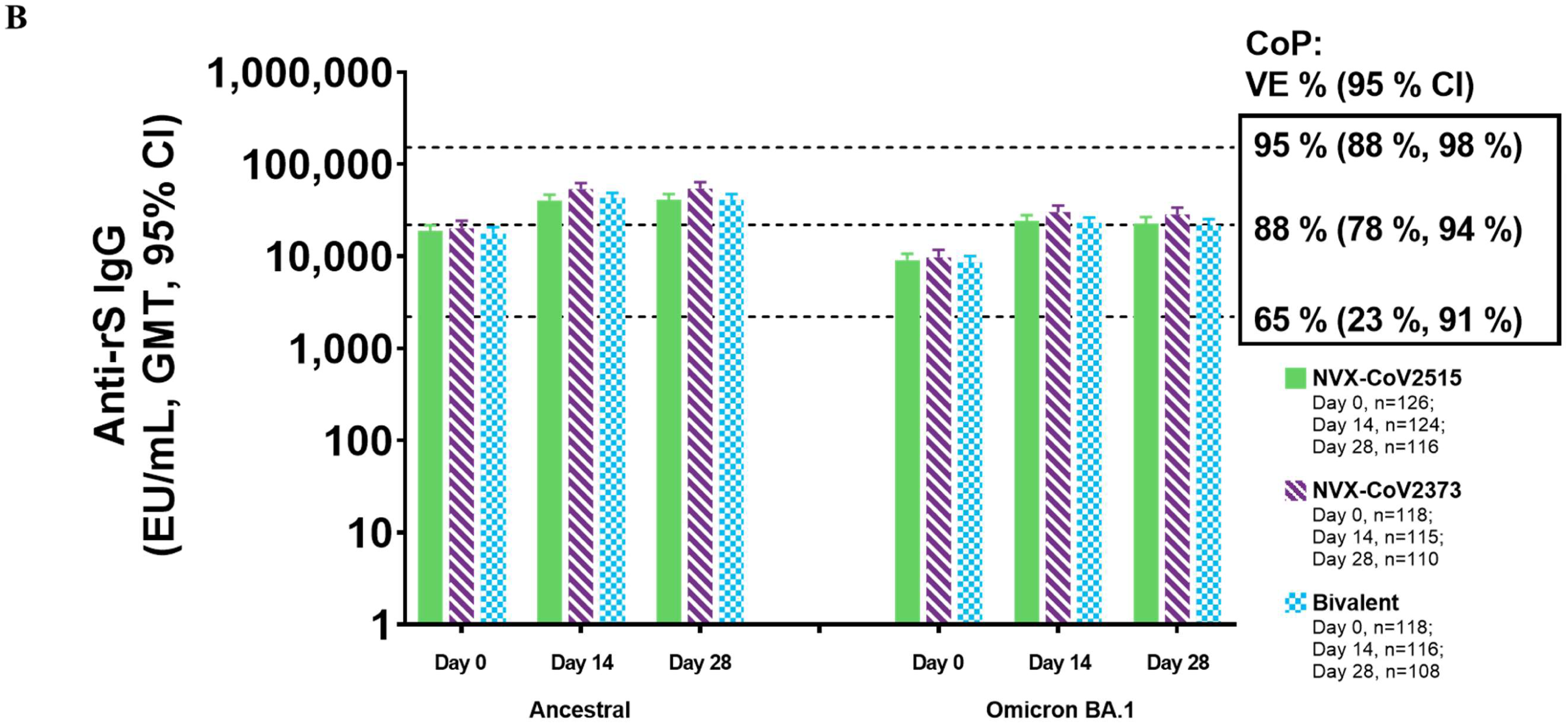
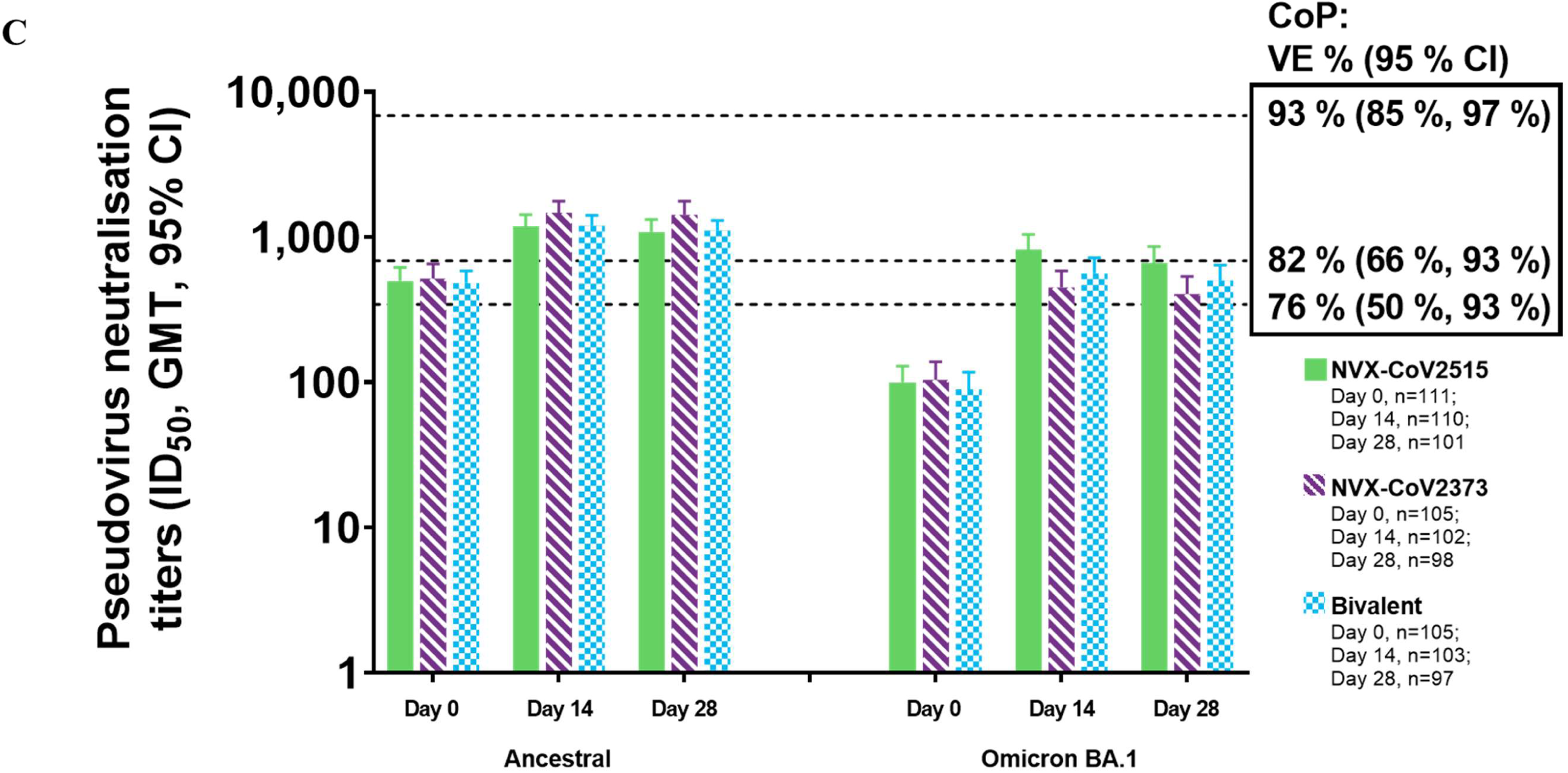
Immunogenicity against ancestral and BA.1 variant strains of SARS-CoV-2, following booster vaccination with NVX-CoV2515, NVX CoV2373, or Bivalent NVX-CoV2373 + NVX-CoV2515 (PP1 Analysis Set). (A) Microneutralization titers for the ancestral and BA.1 variant. (B) Anti-rS IgG concentrations for the ancestral and BA.1 variant. Dotted lines represent approximate correlates of protection (CoP) titers reported in phase 3 studies (**Fong 2023).** (C) Pseudovirus neutralization titers for the ancestral and BA.1 variant. Dotted lines represent approximate CoP titers reported in phase 3 studies (**Fong 2023**).

**Table 2.**
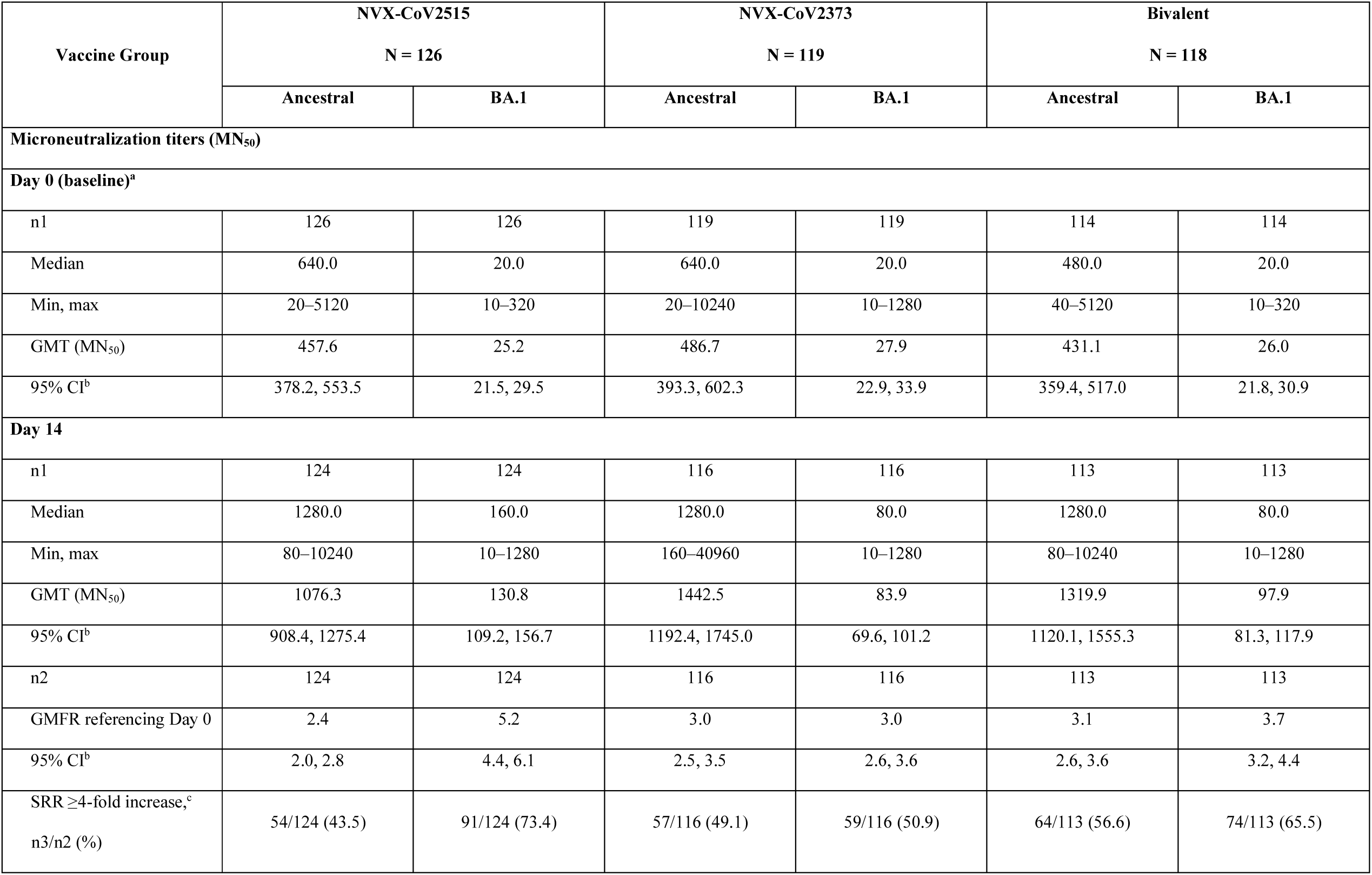
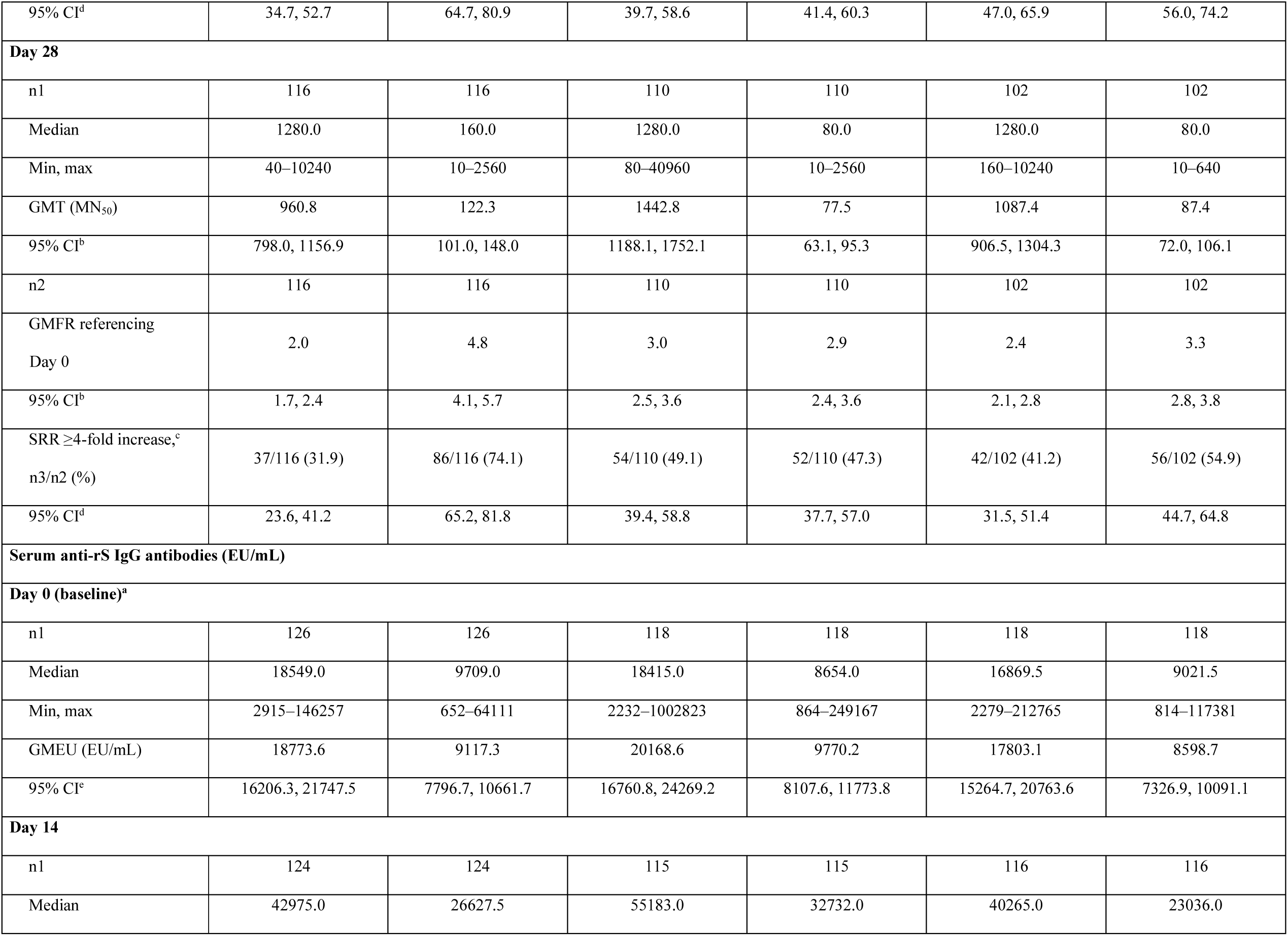
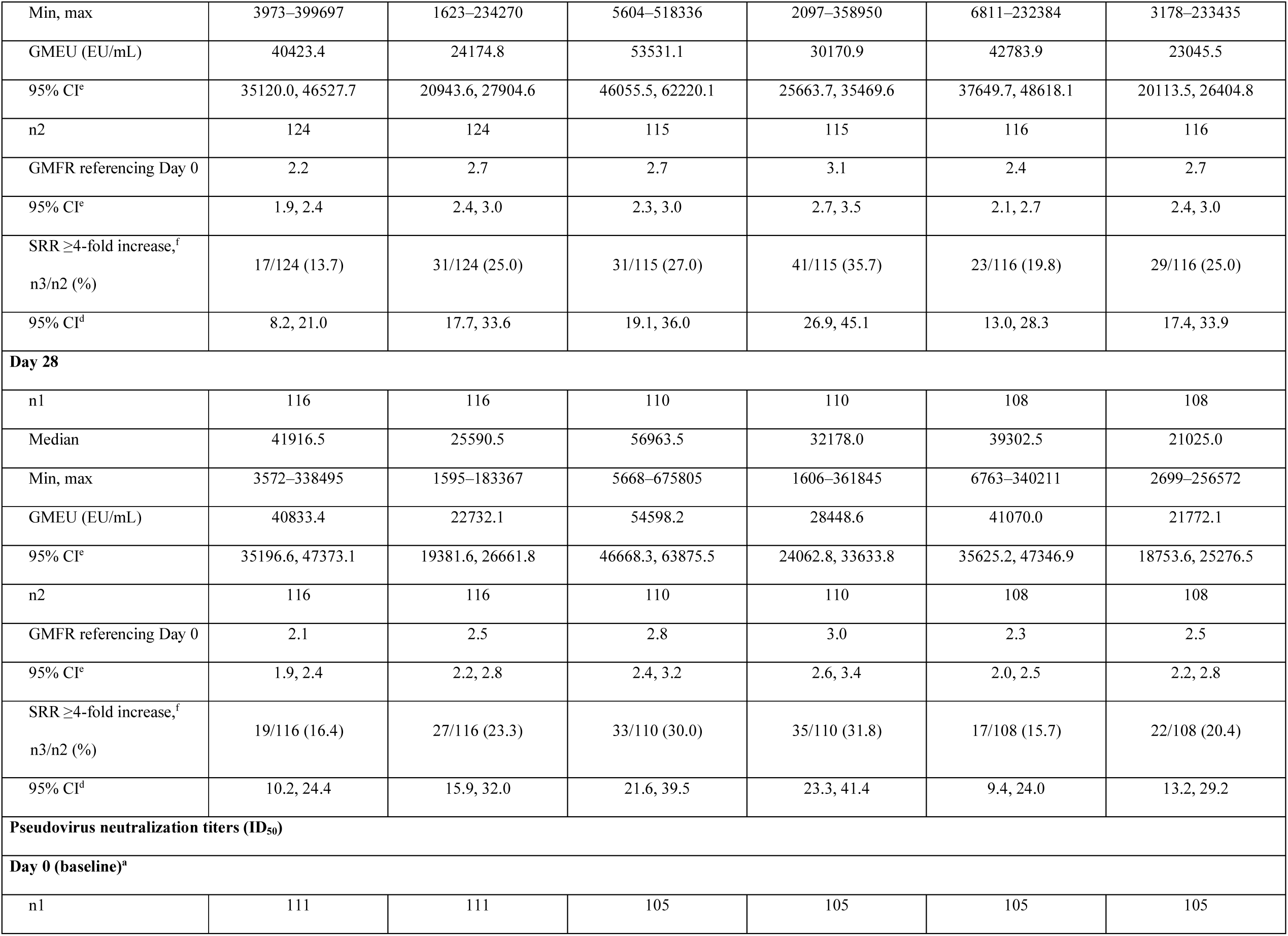
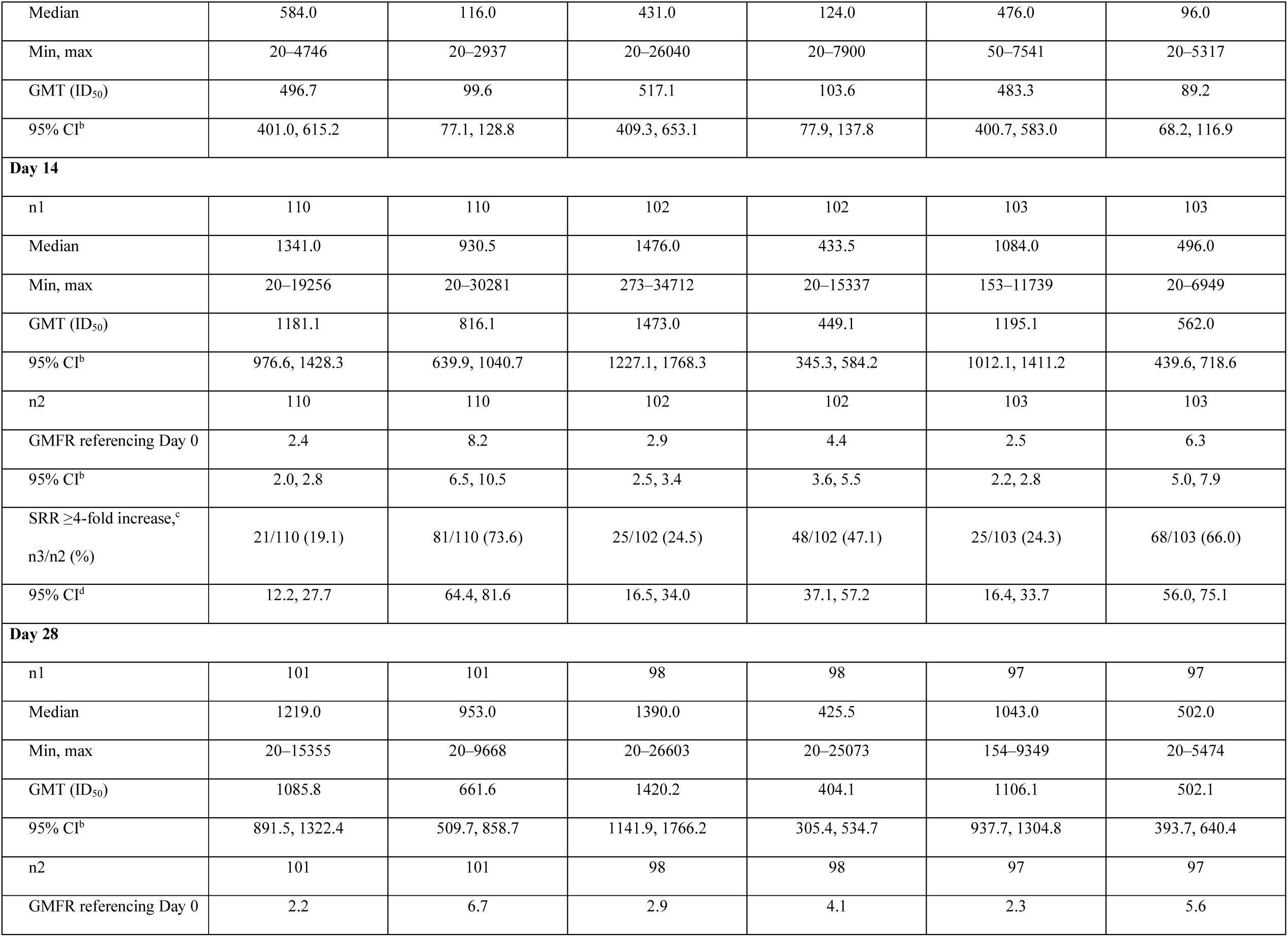
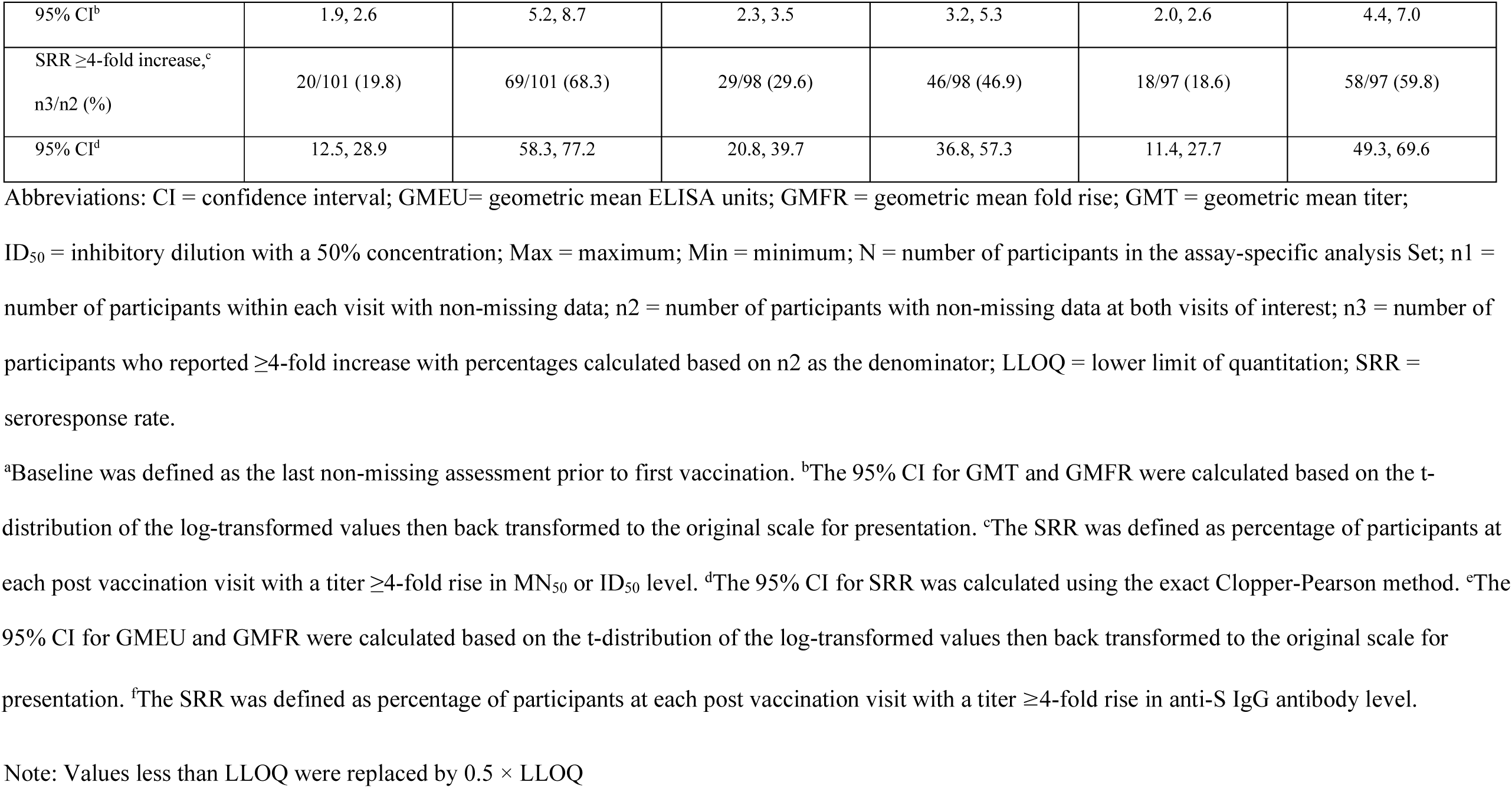
Summary of serum microneutralization titers, anti-rS IgG antibody concentrations, and pseudovirus neutralization titers against the Ancestral and Omicron BA.1 variant strain following a heterologous 4^th^ booster dose (PP1 Analysis Set)

| Vaccine Group | NVX-CoV2515<br>N = 126 |  | NVX-CoV2373<br>N = 119 |  | Bivalent<br>N = 118 |  |
| --- | --- | --- | --- | --- | --- | --- |
|  | Ancestral | BA.1 | Ancestral | BA.1 | Ancestral | BA.1 |
| **Microneutralization titers (MN<sub>50</sub>)** |  |  |  |  |  |  |
| **Day 0 (baseline)<sup>a</sup>** |  |  |  |  |  |  |
| n1 | 126 | 126 | 119 | 119 | 114 | 114 |
| Median | 640.0 | 20.0 | 640.0 | 20.0 | 480.0 | 20.0 |
| Min, max | 20–5120 | 10–320 | 20–10240 | 10–1280 | 40–5120 | 10–320 |
| GMT (MN<sub>50</sub>) | 457.6 | 25.2 | 486.7 | 27.9 | 431.1 | 26.0 |
| 95% CI<sup>b</sup> | 378.2, 553.5 | 21.5, 29.5 | 393.3, 602.3 | 22.9, 33.9 | 359.4, 517.0 | 21.8, 30.9 |
| **Day 14** |  |  |  |  |  |  |
| n1 | 124 | 124 | 116 | 116 | 113 | 113 |
| Median | 1280.0 | 160.0 | 1280.0 | 80.0 | 1280.0 | 80.0 |
| Min, max | 80–10240 | 10–1280 | 160–40960 | 10–1280 | 80–10240 | 10–1280 |
| GMT (MN<sub>50</sub>) | 1076.3 | 130.8 | 1442.5 | 83.9 | 1319.9 | 97.9 |
| 95% CI<sup>b</sup> | 908.4, 1275.4 | 109.2, 156.7 | 1192.4, 1745.0 | 69.6, 101.2 | 1120.1, 1555.3 | 81.3, 117.9 |
| n2 | 124 | 124 | 116 | 116 | 113 | 113 |
| GMFR referencing Day 0 | 2.4 | 5.2 | 3.0 | 3.0 | 3.1 | 3.7 |
| 95% CI<sup>b</sup> | 2.0, 2.8 | 4.4, 6.1 | 2.5, 3.5 | 2.6, 3.6 | 2.6, 3.6 | 3.2, 4.4 |
| SRR ≥4-fold increase,<sup>c</sup><br>n3/n2 (%) | 54/124 (43.5) | 91/124 (73.4) | 57/116 (49.1) | 59/116 (50.9) | 64/113 (56.6) | 74/113 (65.5) |
| 95% CI<sup>d</sup> | 34.7, 52.7 | 64.7, 80.9 | 39.7, 58.6 | 41.4, 60.3 | 47.0, 65.9 | 56.0, 74.2 |
| **Day 28** |  |  |  |  |  |  |
| n1 | 116 | 116 | 110 | 110 | 102 | 102 |
| Median | 1280.0 | 160.0 | 1280.0 | 80.0 | 1280.0 | 80.0 |
| Min, max | 40–10240 | 10–2560 | 80–40960 | 10–2560 | 160–10240 | 10–640 |
| GMT (MN<sub>50</sub>) | 960.8 | 122.3 | 1442.8 | 77.5 | 1087.4 | 87.4 |
| 95% CI<sup>b</sup> | 798.0, 1156.9 | 101.0, 148.0 | 1188.1, 1752.1 | 63.1, 95.3 | 906.5, 1304.3 | 72.0, 106.1 |
| n2 | 116 | 116 | 110 | 110 | 102 | 102 |
| GMFR referencing<br>Day 0 | 2.0 | 4.8 | 3.0 | 2.9 | 2.4 | 3.3 |
| 95% CI<sup>b</sup> | 1.7, 2.4 | 4.1, 5.7 | 2.5, 3.6 | 2.4, 3.6 | 2.1, 2.8 | 2.8, 3.8 |
| SRR ≥4-fold increase,<sup>c</sup><br>n3/n2 (%) | 37/116 (31.9) | 86/116 (74.1) | 54/110 (49.1) | 52/110 (47.3) | 42/102 (41.2) | 56/102 (54.9) |
| 95% CI<sup>d</sup> | 23.6, 41.2 | 65.2, 81.8 | 39.4, 58.8 | 37.7, 57.0 | 31.5, 51.4 | 44.7, 64.8 |
| **Serum anti-rS IgG antibodies (EU/mL)** |  |  |  |  |  |  |
| **Day 0 (baseline)<sup>a</sup>** |  |  |  |  |  |  |
| n1 | 126 | 126 | 118 | 118 | 118 | 118 |
| Median | 18549.0 | 9709.0 | 18415.0 | 8654.0 | 16869.5 | 9021.5 |
| Min, max | 2915–146257 | 652–64111 | 2232–1002823 | 864–249167 | 2279–212765 | 814–117381 |
| GMEU (EU/mL) | 18773.6 | 9117.3 | 20168.6 | 9770.2 | 17803.1 | 8598.7 |
| 95% CI<sup>e</sup> | 16206.3, 21747.5 | 7796.7, 10661.7 | 16760.8, 24269.2 | 8107.6, 11773.8 | 15264.7, 20763.6 | 7326.9, 10091.1 |
| **Day 14** |  |  |  |  |  |  |
| n1 | 124 | 124 | 115 | 115 | 116 | 116 |
| Median | 42975.0 | 26627.5 | 55183.0 | 32732.0 | 40265.0 | 23036.0 |
| Min, max | 3973–399697 | 1623–234270 | 5604–518336 | 2097–358950 | 6811–232384 | 3178–233435 |
| GMEU (EU/mL) | 40423.4 | 24174.8 | 53531.1 | 30170.9 | 42783.9 | 23045.5 |
| 95% CI<sup>e</sup> | 35120.0, 46527.7 | 20943.6, 27904.6 | 46055.5, 62220.1 | 25663.7, 35469.6 | 37649.7, 48618.1 | 20113.5, 26404.8 |
| n2 | 124 | 124 | 115 | 115 | 116 | 116 |
| GMFR referencing Day 0 | 2.2 | 2.7 | 2.7 | 3.1 | 2.4 | 2.7 |
| 95% CI<sup>e</sup> | 1.9, 2.4 | 2.4, 3.0 | 2.3, 3.0 | 2.7, 3.5 | 2.1, 2.7 | 2.4, 3.0 |
| SRR ≥4-fold increase,<sup>f</sup><br>n3/n2 (%) | 17/124 (13.7) | 31/124 (25.0) | 31/115 (27.0) | 41/115 (35.7) | 23/116 (19.8) | 29/116 (25.0) |
| 95% CI<sup>d</sup> | 8.2, 21.0 | 17.7, 33.6 | 19.1, 36.0 | 26.9, 45.1 | 13.0, 28.3 | 17.4, 33.9 |
| **Day 28** |  |  |  |  |  |  |
| n1 | 116 | 116 | 110 | 110 | 108 | 108 |
| Median | 41916.5 | 25590.5 | 56963.5 | 32178.0 | 39302.5 | 21025.0 |
| Min, max | 3572–338495 | 1595–183367 | 5668–675805 | 1606–361845 | 6763–340211 | 2699–256572 |
| GMEU (EU/mL) | 40833.4 | 22732.1 | 54598.2 | 28448.6 | 41070.0 | 21772.1 |
| 95% CI<sup>e</sup> | 35196.6, 47373.1 | 19381.6, 26661.8 | 46668.3, 63875.5 | 24062.8, 33633.8 | 35625.2, 47346.9 | 18753.6, 25276.5 |
| n2 | 116 | 116 | 110 | 110 | 108 | 108 |
| GMFR referencing Day 0 | 2.1 | 2.5 | 2.8 | 3.0 | 2.3 | 2.5 |
| 95% CI<sup>e</sup> | 1.9, 2.4 | 2.2, 2.8 | 2.4, 3.2 | 2.6, 3.4 | 2.0, 2.5 | 2.2, 2.8 |
| SRR ≥4-fold increase,<sup>f</sup><br>n3/n2 (%) | 19/116 (16.4) | 27/116 (23.3) | 33/110 (30.0) | 35/110 (31.8) | 17/108 (15.7) | 22/108 (20.4) |
| 95% CI<sup>d</sup> | 10.2, 24.4 | 15.9, 32.0 | 21.6, 39.5 | 23.3, 41.4 | 9.4, 24.0 | 13.2, 29.2 |
| **Pseudovirus neutralization titers (ID<sub>50</sub>)** |  |  |  |  |  |  |
| **Day 0 (baseline)<sup>a</sup>** |  |  |  |  |  |  |
| n1 | 111 | 111 | 105 | 105 | 105 | 105 |
| Median | 584.0 | 116.0 | 431.0 | 124.0 | 476.0 | 96.0 |
| Min, max | 20–4746 | 20–2937 | 20–26040 | 20–7900 | 50–7541 | 20–5317 |
| GMT (ID<sub>50</sub>) | 496.7 | 99.6 | 517.1 | 103.6 | 483.3 | 89.2 |
| 95% CI<sup>b</sup> | 401.0, 615.2 | 77.1, 128.8 | 409.3, 653.1 | 77.9, 137.8 | 400.7, 583.0 | 68.2, 116.9 |
| **Day 14** |  |  |  |  |  |  |
| n1 | 110 | 110 | 102 | 102 | 103 | 103 |
| Median | 1341.0 | 930.5 | 1476.0 | 433.5 | 1084.0 | 496.0 |
| Min, max | 20–19256 | 20–30281 | 273–34712 | 20–15337 | 153–11739 | 20–6949 |
| GMT (ID<sub>50</sub>) | 1181.1 | 816.1 | 1473.0 | 449.1 | 1195.1 | 562.0 |
| 95% CI<sup>b</sup> | 976.6, 1428.3 | 639.9, 1040.7 | 1227.1, 1768.3 | 345.3, 584.2 | 1012.1, 1411.2 | 439.6, 718.6 |
| n2 | 110 | 110 | 102 | 102 | 103 | 103 |
| GMFR referencing Day 0 | 2.4 | 8.2 | 2.9 | 4.4 | 2.5 | 6.3 |
| 95% CI<sup>b</sup> | 2.0, 2.8 | 6.5, 10.5 | 2.5, 3.4 | 3.6, 5.5 | 2.2, 2.8 | 5.0, 7.9 |
| SRR ≥4-fold increase,<sup>c</sup><br>n3/n2 (%) | 21/110 (19.1) | 81/110 (73.6) | 25/102 (24.5) | 48/102 (47.1) | 25/103 (24.3) | 68/103 (66.0) |
| 95% CI<sup>d</sup> | 12.2, 27.7 | 64.4, 81.6 | 16.5, 34.0 | 37.1, 57.2 | 16.4, 33.7 | 56.0, 75.1 |
| **Day 28** |  |  |  |  |  |  |
| n1 | 101 | 101 | 98 | 98 | 97 | 97 |
| Median | 1219.0 | 953.0 | 1390.0 | 425.5 | 1043.0 | 502.0 |
| Min, max | 20–15355 | 20–9668 | 20–26603 | 20–25073 | 154–9349 | 20–5474 |
| GMT (ID<sub>50</sub>) | 1085.8 | 661.6 | 1420.2 | 404.1 | 1106.1 | 502.1 |
| 95% CI<sup>b</sup> | 891.5, 1322.4 | 509.7, 858.7 | 1141.9, 1766.2 | 305.4, 534.7 | 937.7, 1304.8 | 393.7, 640.4 |
| n2 | 101 | 101 | 98 | 98 | 97 | 97 |
| GMFR referencing Day 0 | 2.2 | 6.7 | 2.9 | 4.1 | 2.3 | 5.6 |
| 95% CI<sup>b</sup> | 1.9, 2.6 | 5.2, 8.7 | 2.3, 3.5 | 3.2, 5.3 | 2.0, 2.6 | 4.4, 7.0 |
| SRR ≥4-fold increase,<sup>c</sup><br>n3/n2 (%) | 20/101 (19.8) | 69/101 (68.3) | 29/98 (29.6) | 46/98 (46.9) | 18/97 (18.6) | 58/97 (59.8) |
| 95% CI<sup>d</sup> | 12.5, 28.9 | 58.3, 77.2 | 20.8, 39.7 | 36.8, 57.3 | 11.4, 27.7 | 49.3, 69.6 |

Abbreviations: CI = confidence interval; GMEU= geometric mean ELISA units; GMFR = geometric mean fold rise; GMT = geometric mean titer;
ID<sub>50</sub> = inhibitory dilution with a 50% concentration; Max = maximum; Min = minimum; N = number of participants in the assay-specific analysis Set; n1 = number of participants within each visit with non-missing data; n2 = number of participants with non-missing data at both visits of interest; n3 = number of participants who reported ≥ 4-fold increase with percentages calculated based on n2 as the denominator; LLOQ = lower limit of quantitation; SRR = seroresponse rate.
<sup>a</sup>Baseline was defined as the last non-missing assessment prior to first vaccination. <sup>b</sup>The 95% CI for GMT and GMFR were calculated based on the t-distribution of the log-transformed values then back transformed to the original scale for presentation. <sup>c</sup>The SRR was defined as percentage of participants at each post vaccination visit with a titer ≥ 4-fold rise in MN<sub>50</sub> or ID<sub>50</sub> level. <sup>d</sup>The 95% CI for SRR was calculated using the exact Clopper-Pearson method. <sup>e</sup>The 95% CI for GMEU and GMFR were calculated based on the t-distribution of the log-transformed values then back transformed to the original scale for presentation. <sup>f</sup>The SRR was defined as percentage of participants at each post vaccination visit with a titer ≥ 4-fold rise in anti-S IgG antibody level.
Note: Values less than LLOQ were replaced by $0.5 \times \text{LLOQ}$

Anti-spike IgG assay data indicate that the highest concentrations of Omicron BA.1 IgG are achieved with NVX-CoV2373, highlighting the vaccine’s cross-reactive nature with the Omicron BA.1 sublineage. Anti-spike IgG antibody levels (geometric mean ELISA units, GMEU) [95% CI] for the BA.1 assay were 30,170.9 [25,663.7, 35,469.6], 24,174.8 [20,943.6, 27,904.6], and 23,045.5 [20,113.5, 26,404.8] EU/mL for NVX-CoV2373, NVX-CoV2515, and the bivalent vaccine, respectively (**Figure 1**). A somewhat similar pattern of response was seen using the ancestral strain anti-spike IgG assay, with GMEUs of 53,531.1 [46,055.5, 62,220.1], 40,423.4 [35,120.0, 46,527.7] and 42,783.9 [37,649.7, 48,618.1] EU/mL for NVX-CoV2373, NVX-CoV2515, and the bivalent vaccine, respectively (**Figure 1**).

Pseudovirus neutralization GMTs against the Omicron BA.1 sublineage were the highest in participants receiving NVX-CoV2515, followed by those receiving the bivalent vaccine and NVX-CoV2373 (ID50 titers [95% CI] of 816.1 [639.9, 1040.7], 562.0 [439.6, 718.6], and 449.1 [345.3, 584.2], respectively) (**Figure 1**). ID50 GMTs for the ancestral strain pseudovirus neutralization assay were somewhat lower in participants receiving NVX-CoV2515 and the bivalent vaccine compared with NVX-CoV2373 (ID50 titers [95% CI] of 1181.1 [976.6, 1428.3], 1195.1 [1012.1, 1411.2], and 1473.0 [1227.1, 1768.3], respectively) (**Table 2**).

### Per Protocol Analysis Set 2

Microneutralization assay GMTs against the Omicron BA.1 sublineage for participants in the PP2 population, which included those with a positive baseline SARS-CoV-2 result, were the highest in participants receiving NVX-CoV2515, followed by those receiving the bivalent vaccine and NVX-CoV2373 (MN50 titers [95% CI] of 318.2 [269.8, 375.3], 252.7 [213.1, 299.7], and 218.1 [186.0, 255.7], respectively) (**Figure 2**). NVX-CoV2515 induced a non-inferior SRR against the Omicron BA.1 subvariant virus versus NVX-CoV2373 (54.3% [134/247] vs 32.0% [78/244]) at Day 14, with similar results on Day 28 (**Table 3**). MN50 GMTs for the ancestral strain microneutralization assay were somewhat lower in participants receiving NVX-CoV2515 compared with the bivalent and NVX-CoV2373 vaccines (MN50 titers [95% CI] of 2206.2 [1910.0, 2548.4], 2544.7 [2194.5, 2950.9], and 2702.0 [2347.9, 3109.4], respectively) (**Table 3**).

**Figure 2.**
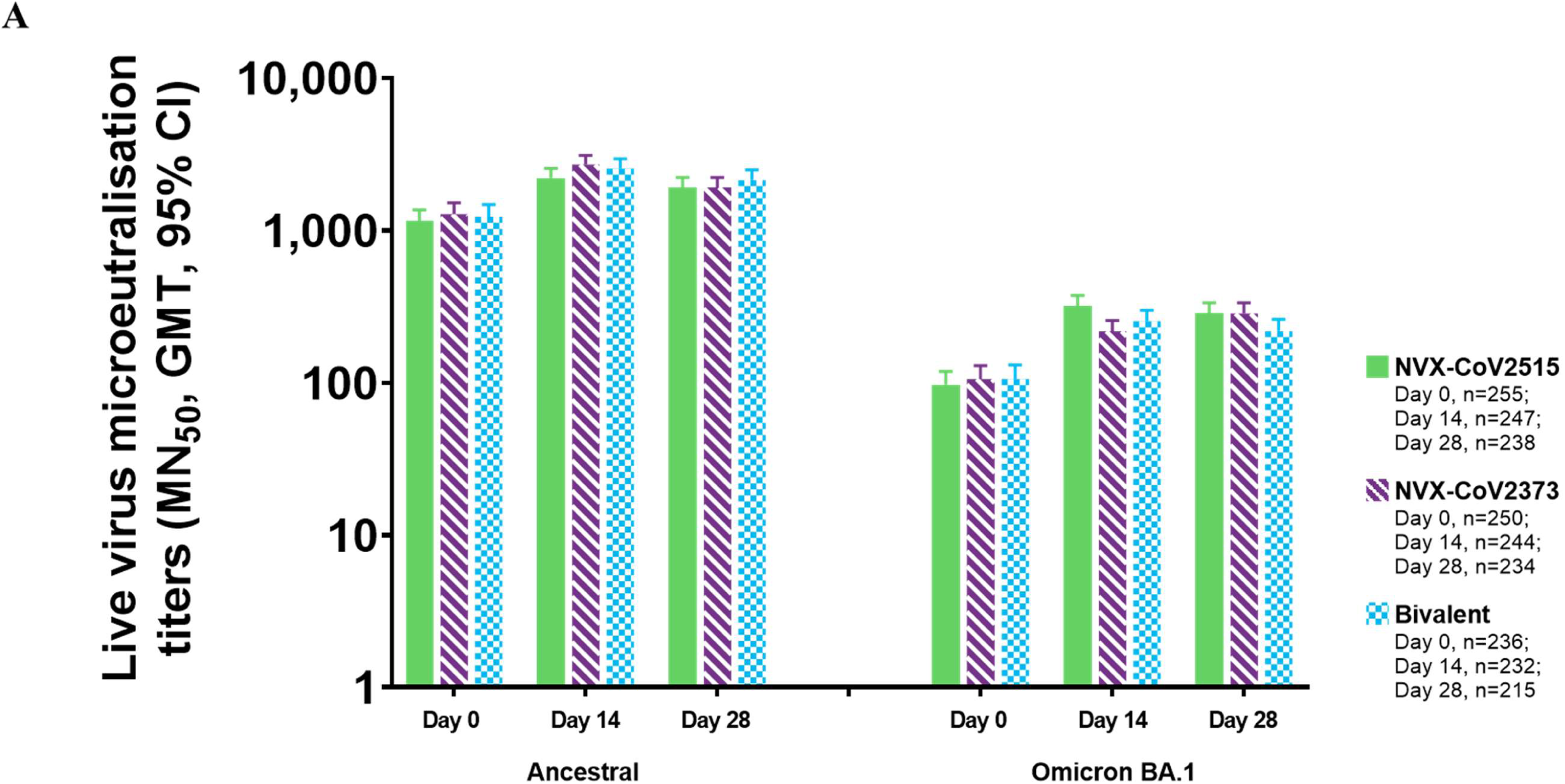
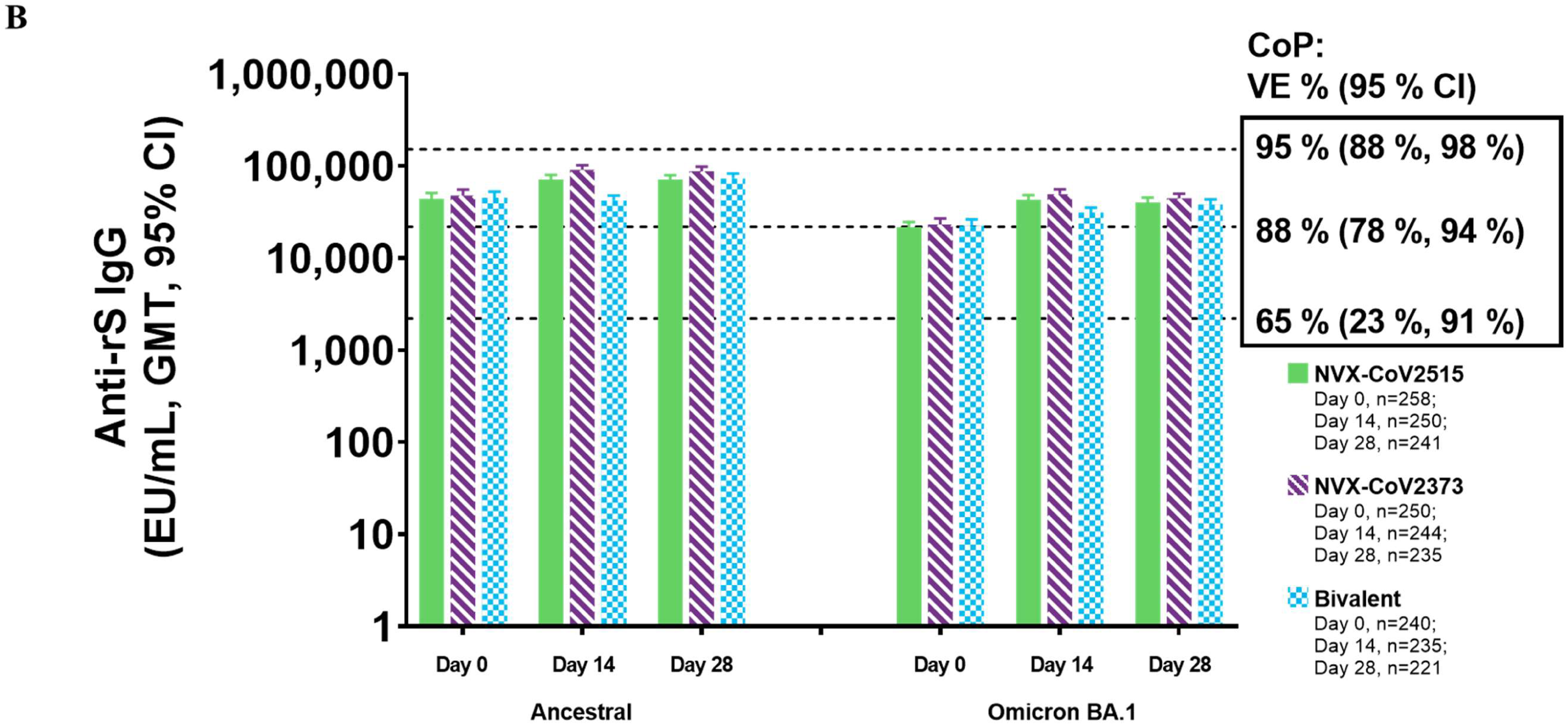
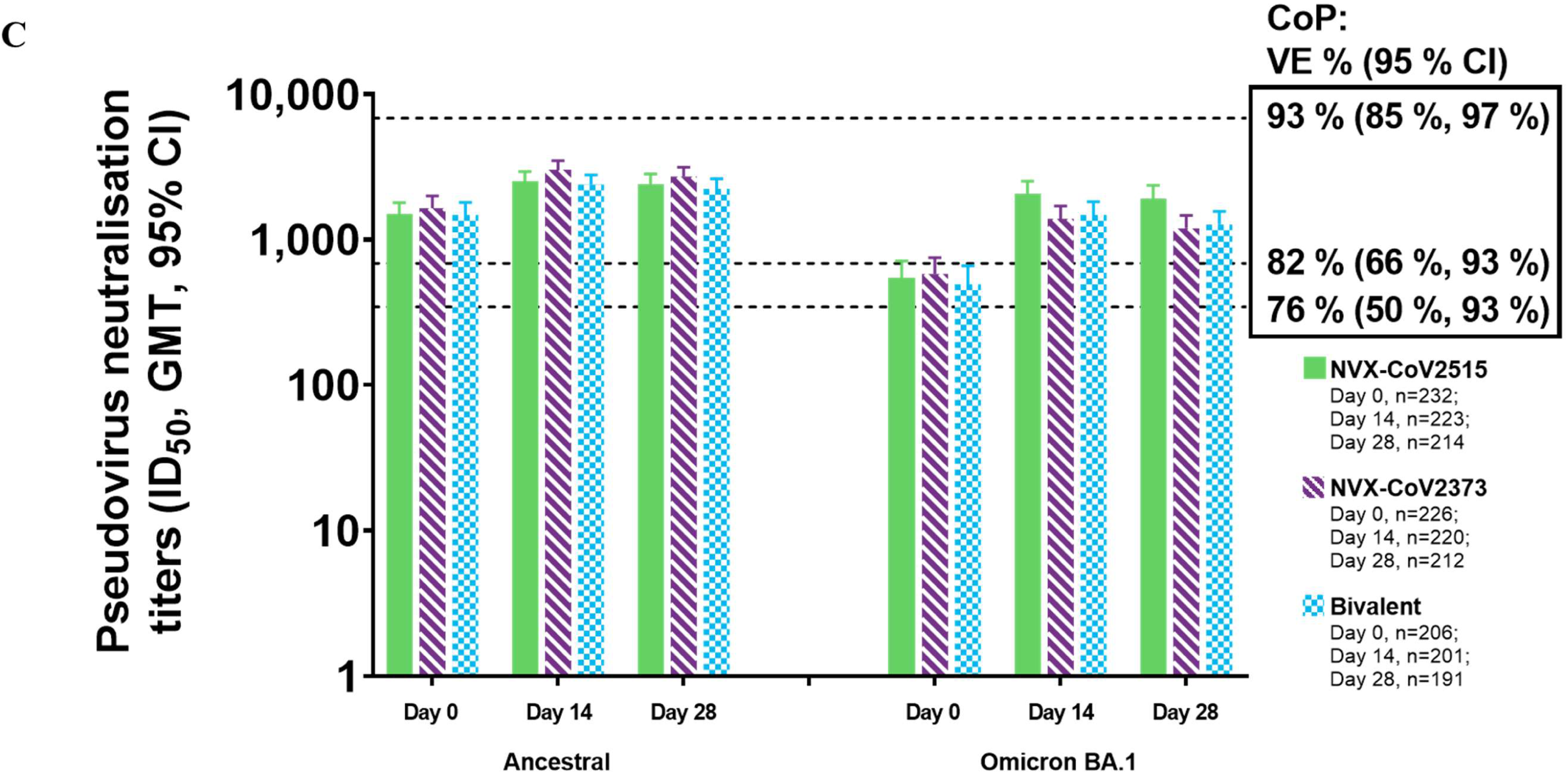
Immunogenicity against ancestral and BA.1 variant strains of SARS-CoV-2, following booster vaccination with NVX-CoV2515, NVX CoV2373, or Bivalent NVX-CoV2373 + NVX-CoV2515 (PP2 Analysis Set). (A) Microneutralization titers for the ancestral and BA.1 variant. (B) Anti-rS IgG concentrations for the ancestral and BA.1 variant. Dotted lines represent approximate CoP titers reported in phase 3 studies (**Fong 2023**). (C) Pseudovirus neutralization titers for the ancestral and BA.1 variant. Dotted lines represent approximate CoP titers reported in phase 3 studies (**Fong 2023**).

**Table 3.** Summary of serum microneutralization titers, anti-rS IgG antibody concentrations, and pseudovirus neutralization titers against the Ancestral and Omicron BA.1 variant strain following a heterologous 4^th^ booster dose (PP2 Analysis Set)

| Vaccine Group | NVX-CoV2515<br>N = 286 |  | NVX-CoV2373<br>N = 274 |  | Bivalent<br>N = 269 |  |
| --- | --- | --- | --- | --- | --- | --- |
|  | Ancestral | BA.1 | Ancestral | BA.1 | Ancestral | BA.1 |
| **Microneutralization titers (MN<sub>50</sub>)** |  |  |  |  |  |  |
| **Day 0 (baseline)<sup>a</sup>** |  |  |  |  |  |  |
| n1 | 255 | 255 | 250 | 250 | 236 | 236 |
| Median | 1280.0 | 160.0 | 1280.0 | 160.0 | 1280.0 | 160.0 |
| Min–max | 20–40960 | 10–2560 | 20–40960 | 10–2560 | 40–81920 | 10–10240 |
| GMT (MN<sub>50</sub>) | 1151.3 | 97.3 | 1280.0 | 105.9 | 1232.0 | 106.1 |
| 95% CI<sup>b</sup> | 973.6, 1361.4 | 79.6, 118.9 | 1078.1, 1519.7 | 86.4, 129.7 | 1026.0, 1479.5 | 85.8, 131.1 |
| **Day 14** |  |  |  |  |  |  |
| n1 | 247 | 247 | 244 | 244 | 232 | 232 |
| Median | 2560.0 | 320.0 | 2560.0 | 320.0 | 2560.0 | 320.0 |
| Min–max | 80–40960 | 10–5120 | 160–81920 | 10–2560 | 80–163840 | 10–5120 |
| GMT (MN<sub>50</sub>) | 2206.2 | 318.2 | 2702.0 | 218.1 | 2544.7 | 252.7 |
| 95% CI<sup>b</sup> | 1910.0, 2548.4 | 269.8, 375.3 | 2347.9, 3109.4 | 186.0, 255.7 | 2194.5, 2950.9 | 213.1, 299.7 |
| n2 | 247 | 247 | 244 | 244 | 232 | 232 |
| GMFR referencing Day 0 | 1.9 | 3.3 | 2.1 | 2.1 | 2.1 | 2.4 |
| 95% CI<sup>b</sup> | 1.8, 2.1 | 2.9, 3.7 | 1.9, 2.4 | 1.8, 2.3 | 1.9, 2.3 | 2.1, 2.7 |
| SRR ≥4-fold increase,<sup>c</sup><br>n3/n2 (%) | 79/247 (32.0) | 134/247 (54.3) | 80/244 (32.8) | 78/244 (32.0) | 83/232 (35.8) | 95/232 (40.9) |
| 95% CI<sup>d</sup> | 26.2, 38.2 | 47.8, 60.6 | 26.9, 39.1 | 26.2, 38.2 | 29.6, 42.3 | 34.6, 47.6 |
| **Day 28** |  |  |  |  |  |  |
| n1 | 238 | 238 | 234 | 234 | 215 | 215 |
| Median | 2560.0 | 320.0 | 2560.0 | 160.0 | 2560.0 | 160.0 |
| Min–max | 40–20480 | 10–5120 | 80–40960 | 10–2560 | 160–81920 | 10–10240 |
| GMT (MN<sub>50</sub>) | 1918.8 | 284.8 | 2456.0 | 195.7 | 2144.0 | 218.7 |
| 95% CI<sup>b</sup> | 1657.9, 2220.6 | 241.8, 335.4 | 2145.2, 2811.8 | 165.7, 231.2 | 1842.3, 2495.2 | 183.1, 261.3 |
| n2 | 238 | 238 | 234 | 234 | 215 | 215 |
| GMFR referencing Day 0 | 1.6 | 2.9 | 1.9 | 1.9 | 1.7 | 2.0 |
| 95% CI<sup>b</sup> | 1.5, 1.8 | 2.5, 3.3 | 1.7, 2.2 | 1.6, 2.2 | 1.5, 1.9 | 1.8, 2.2 |
| SRR ≥4-fold increase,<sup>c</sup><br>n3/n2 (%) | 56/238 (23.5) | 125/238 (52.5) | 68/234 (29.1) | 65/234 (27.8) | 59/215 (27.4) | 72/215 (33.5) |
| 95% CI<sup>d</sup> | 18.3, 29.4 | 46.0, 59.0 | 23.3, 35.3 | 22.1, 34.0 | 21.6, 33.9 | 27.2, 40.2 |
| **Serum anti-rS IgG antibodies (EU/mL)** |  |  |  |  |  |  |
| **Day 0 (baseline)<sup>a</sup>** |  |  |  |  |  |  |
| n1 | 258 | 258 | 250 | 250 | 240 | 240 |
| Median | 45426.5 | 22876.0 | 54982.0 | 28832.0 | 49528.0 | 25602.0 |
| Min–max | 2915–576831 | 652–353367 | 2232–1002823 | 864–254105 | 2279–2034472 | 814–1121904 |
| GMEU (EU/mL) | 44166.7 | 21446.9 | 47810.6 | 23215.7 | 45047.6 | 22400.7 |
| 95% CI<sup>e</sup> | 38369.6, 50839.6 | 18570.6, 24768.7 | 41184.3, 55503.1 | 19953.4, 27011.3 | 38572.8, 52609.1 | 19088.9, 26287.2 |
| **Day 14** |  |  |  |  |  |  |
| n1 | 250 | 250 | 244 | 244 | 235 | 235 |
| Median | 72765.5 | 41964.5 | 95097.0 | 50675.0 | 79657.0 | 43289.0 |
| Min–max | 3973–894339 | 1623–813679 | 5604–822744 | 2097–395875 | 6811–1656933 | 3178–1074179 |
| GMEU (EU/mL) | 71076.4 | 42835.5 | 90962.2 | 49727.7 | 78191.9 | 42462.1 |
| 95% CI<sup>e</sup> | 63012.1, 80172.9 | 37883.8, 48434.4 | 81361.1, 101696.2 | 44331.1, 55781.1 | 69489.1, 87984.5 | 37628.9, 47916.2 |
| n2 | 250 | 250 | 244 | 244 | 235 | 235 |
| GMFR referencing Day 0 | 1.6 | 2.0 | 1.9 | 2.1 | 1.7 | 1.9 |
| 95% CI<sup>e</sup> | 1.5, 1.7 | 1.8, 2.2 | 1.8, 2.1 | 2.0, 2.3 | 1.6, 1.9 | 1.7, 2.0 |
| SRR ≥4-fold increase,<sup>f</sup><br>n3/n2 (%) | 18/250 (7.2) | 34/250 (13.6) | 35/244 (14.3) | 42/244 (17.2) | 23/235 (9.8) | 29/235 (12.3) |
| 95% CI<sup>d</sup> | 4.3, 11.1 | 9.6, 18.5 | 10.2, 19.4 | 12.7, 22.5 | 6.3, 14.3 | 8.4, 17.2 |
| **Day 28** |  |  |  |  |  |  |
| n1 | 241 | 241 | 235 | 235 | 221 | 221 |
| Median | 72699.0 | 42388.0 | 92903.0 | 44971.0 | 70184.0 | 36642.0 |
| Min–max | 3572–755671 | 1595–543014 | 5668–675805 | 1606–375634 | 6763–2299922 | 2699–1217244 |
| GMC (EU/mL) | 70523.9 | 40044.9 | 87969.9 | 44756.4 | 73321.3 | 38315.6 |
| 95% CI<sup>e</sup> | 62487.3, 79594.2 | 35308.6, 45416.6 | 78701.3, 98330.0 | 39966.0, 50120.9 | 64771.3, 83000.0 | 33783.4, 43455.9 |
| n2 | 241 | 241 | 235 | 235 | 221 | 221 |
| GMFR referencing Day 0 | 1.6 | 1.8 | 1.9 | 2.0 | 1.6 | 1.7 |
| 95% CI<sup>e</sup> | 1.4, 1.7 | 1.7, 2.0 | 1.7, 2.0 | 1.8, 2.1 | 1.5, 1.8 | 1.6, 1.8 |
| SRR ≥4-fold increase,<sup>f</sup><br>n3/n2 (%) | 19/241 (7.9) | 28/241 (11.6) | 34/235 (14.5) | 37/235 (15.7) | 17/221 (7.7) | 22/221 (10.0) |
| 95% CI<sup>d</sup> | 4.8, 12.0 | 7.9, 16.4 | 10.2, 19.6 | 11.3, 21.0 | 4.5, 12.0 | 6.3, 14.7 |
| **Pseudovirus neutralization titers (ID<sub>50</sub>)** |  |  |  |  |  |  |
| **Day 0 (baseline)<sup>a</sup>** |  |  |  |  |  |  |
| n1 | 232 | 232 | 226 | 226 | 206 | 206 |
| Median | 1788.5 | 808.0 | 2224.5 | 962.0 | 1425.5 | 623.5 |
| Min, max | 20–39721 | 20–30397 | 20–28737 | 20–18494 | 50–81399 | 20–84031 |
| GMT (ID<sub>50</sub>) | 1491.2 | 548.4 | 1646.5 | 579.2 | 1476.4 | 491.9 |
| 95% CI<sup>b</sup> | 1237.3, 1797.1 | 421.7, 713.2 | 1357.6, 1996.8 | 445.6, 752.8 | 1208.2, 1804.1 | 366.8, 659.6 |
| **Day 14** |  |  |  |  |  |  |
| n1 | 223 | 223 | 220 | 220 | 201 | 201 |
| Median | 2464.0 | 2202.0 | 3121.5 | 1690.5 | 2526.0 | 1677.0 |
| Min, max | 20–34131 | 20–41483 | 273–34859 | 20–23694 | 153–71353 | 20–72247 |
| GMT (ID<sub>50</sub>) | 2514.9 | 2071.5 | 3027.9 | 1390.3 | 2390.1 | 1480.4 |
| 95% CI<sup>b</sup> | 2149.8, 2942.0 | 1701.1, 2522.5 | 2625.7, 3491.6 | 1134.6, 1703.8 | 2050.2, 2786.3 | 1202.8, 1822.1 |
| n2 | 223 | 223 | 220 | 220 | 201 | 201 |
| GMFR referencing Day 0 | 1.7 | 3.9 | 1.8 | 2.4 | 1.6 | 3.0 |
| 95% CI<sup>b</sup> | 1.5, 1.9 | 3.3, 4.6 | 1.7, 2.0 | 2.1, 2.8 | 1.5, 1.8 | 2.5, 3.5 |
| SRR ≥4-fold increase,<sup>c</sup><br>n3/n2 (%) | 25/223 (11.2) | 102/223 (45.7) | 26/220 (11.8) | 53/220 (24.1) | 25/201 (12.4) | 71/201 (35.3) |
| 95% CI<sup>d</sup> | 7.4, 16.1 | 39.1, 52.5 | 7.9, 16.8 | 18.6, 30.3 | 8.2, 17.8 | 28.7, 42.4 |
| **Day 28** |  |  |  |  |  |  |
| n1 | 214 | 214 | 212 | 212 | 191 | 191 |
| Median | 2445.0 | 2127.5 | 2875.0 | 1332.5 | 2193.0 | 1303.0 |
| Min, max | 20–45309 | 20–70894 | 20–31723 | 20–25073 | 154–168338 | 20–95088 |
| GMT (ID<sub>50</sub>) | 2411.6 | 1918.0 | 2716.9 | 1195.7 | 2236.0 | 1272.4 |
| 95% CI<sup>b</sup> | 2055.6, 2829.2 | 1558.2, 2361.0 | 2340.0, 3154.5 | 974.9, 1466.6 | 1909.8, 2617.9 | 1033.5, 1566.6 |
| n2 | 214 | 214 | 212 | 212 | 191 | 191 |
| GMFR referencing Day 0 | 1.6 | 3.4 | 1.7 | 2.1 | 1.5 | 2.6 |
| 95% CI<sup>b</sup> | 1.5, 1.8 | 2.9, 4.0 | 1.5, 1.9 | 1.8, 2.5 | 1.4, 1.7 | 2.2, 3.0 |
| SRR ≥4-fold increase,<sup>c</sup><br>n3/n2 (%) | 22/214 (10.3) | 84/214 (39.3) | 29/212 (13.7) | 48/212 (22.6) | 19/191 (9.9) | 61/191 (31.9) |
| 95% CI<sup>d</sup> | 6.6, 15.2 | 32.7, 46.1 | 9.4, 19.1 | 17.2, 28.9 | 6.1, 15.1 | 25.4, 39.1 |

Abbreviations: CI = confidence interval; GMEU = geometric mean ELISA units; GMFR = geometric mean fold rise; GMT = geometric mean titer;
ID<sub>50</sub> = inhibitory dilution with a 50% concentration; Max = maximum; Min = minimum; N = number of participants in the assay-specific analysis Set; n1 = number of participants within each visit with non-missing data; n2 = number of participants with non-missing data at both visits of interest; n3 = number of participants who reported ≥ 4-fold increase with percentages calculated based on n2 as the denominator; LLOQ = lower limit of quantitation; SRR = seroresponse rate.
<sup>a</sup>Baseline was defined as the last non-missing assessment prior to first vaccination. <sup>b</sup>The 95% CI for GMT and GMFR were calculated based on the t-distribution of the log-transformed values then back transformed to the original scale for presentation. <sup>c</sup>The SRR was defined as percentage of participants at each post vaccination visit with a titer ≥ 4-fold rise in MN<sub>50</sub> or ID<sub>50</sub> level. <sup>d</sup>The 95% CI for SRR was calculated using the exact Clopper-Pearson method. <sup>e</sup>The 95% CI for GMEU and GMFR were calculated based on the t-distribution of the log-transformed values then back transformed to the original scale for presentation. <sup>f</sup>The SRR was defined as percentage of participants at each post vaccination visit with a titer ≥ 4-fold rise in anti-S IgG antibody level.
Note: Values less than LLOQ were replaced by $0.5 \times \text{LLOQ}$

Similar to the PP1 population, anti-spike IgG assay data indicate the highest levels of BA.1 IgG are achieved with vaccination using NVX-CoV2373, indicating cross-reactivity with the Omicron BA.1 sublineage (**Table 3**). Anti-spike IgG GMEUs [95% CI] for the BA.1 assay were 49,727.7 [44,331.1, 55,781.1], 42,835.5 [37,883.8, 48,434.4], and 42,462.1 [37,628.9, 47,916.2]

EU/mL for NVX-CoV2373, NVX-CoV2515, and the bivalent vaccine, respectively (**Figure 2**). A relatively similar pattern of response was seen using the ancestral strain anti-spike IgG assay, with GMEUs of 90,962.2 [81,361.1, 101,696.2], 78,191.9 [69,489.1, 87,984.5], and 71,076.4 [63,012.1, 80,172.9] EU/mL for NVX-CoV2373, bivalent vaccine, and NVX-CoV2515, respectively (**Figure 2**).

### Safety Analysis Set

The overall rates of solicited local and systemic reactions reported within 7 days after booster vaccination were similar across all 3 investigational products (**Figure 3**). Rates of solicited local events of any grade were 69.3% (196/286; Grade 3+: 1.8% [5/286]), 71.0% (193/274; 0.4% [1/274]), and 64.6% (173/269; 1.1% [3/269] for NVX-CoV2515, NVX-CoV2373, and the bivalent vaccine, respectively). No Grade 4 solicited local treatment-emergent adverse events (TEAEs) were reported. Local reactions were generally short-lived, with a median duration of 1.0 day for all events except tenderness (2.0 days). Solicited systemic reactions were similar across the groups with event rates [Grade 3+ rates] of 62.2% [7.3%], 58.1% [3.7%], and 61.9% [3.0%] for NVX-CoV2515, NVX-CoV2373, and the bivalent vaccine, respectively. There was one Grade 4 solicited systemic TEAE (fever) reported in the NVX-CoV2515 group. Solicited systemic reactions were transient in nature, with a median duration of 1.0 day for all events except fatigue, which had a median duration of 2.0 days.

**Figure 3.**
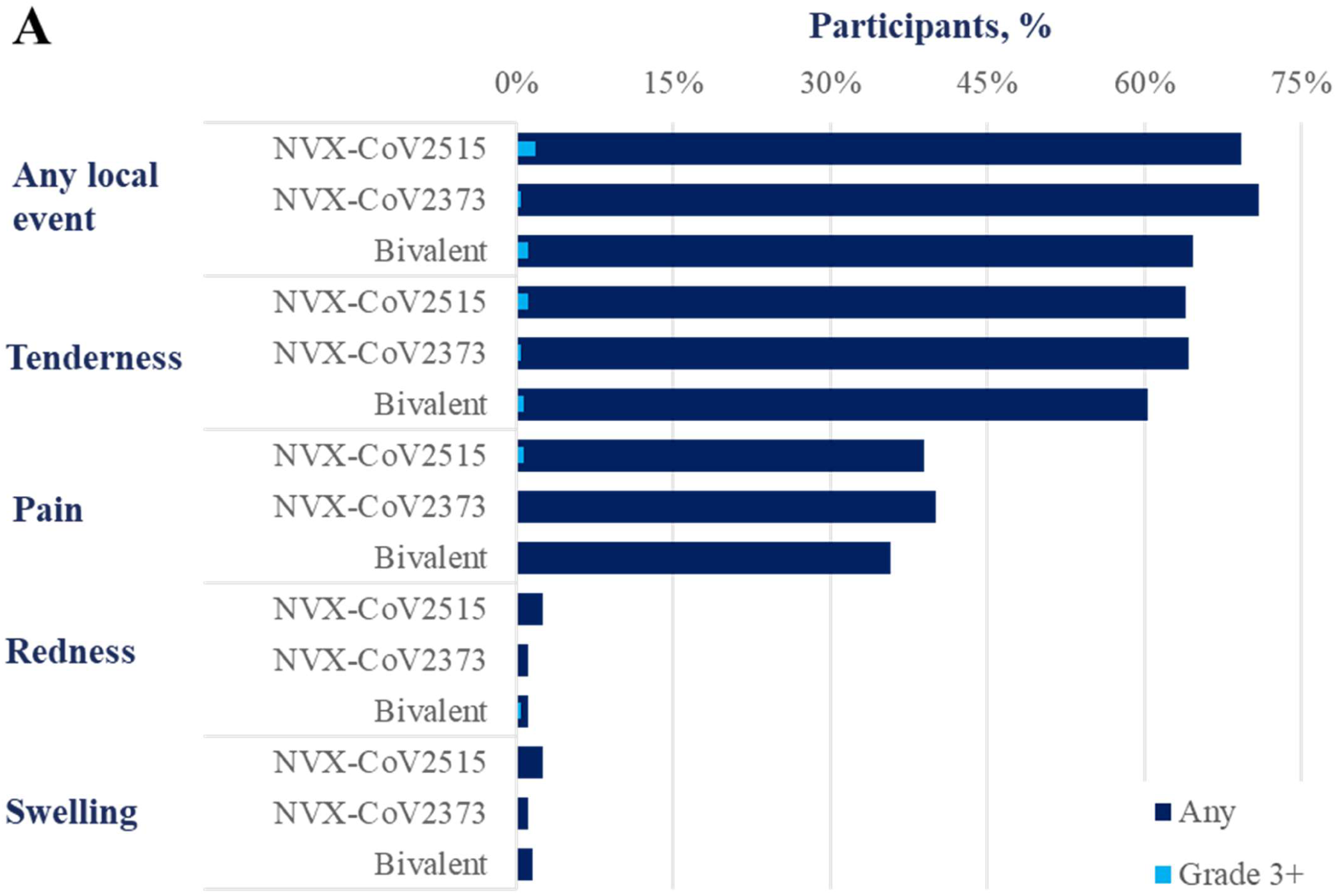
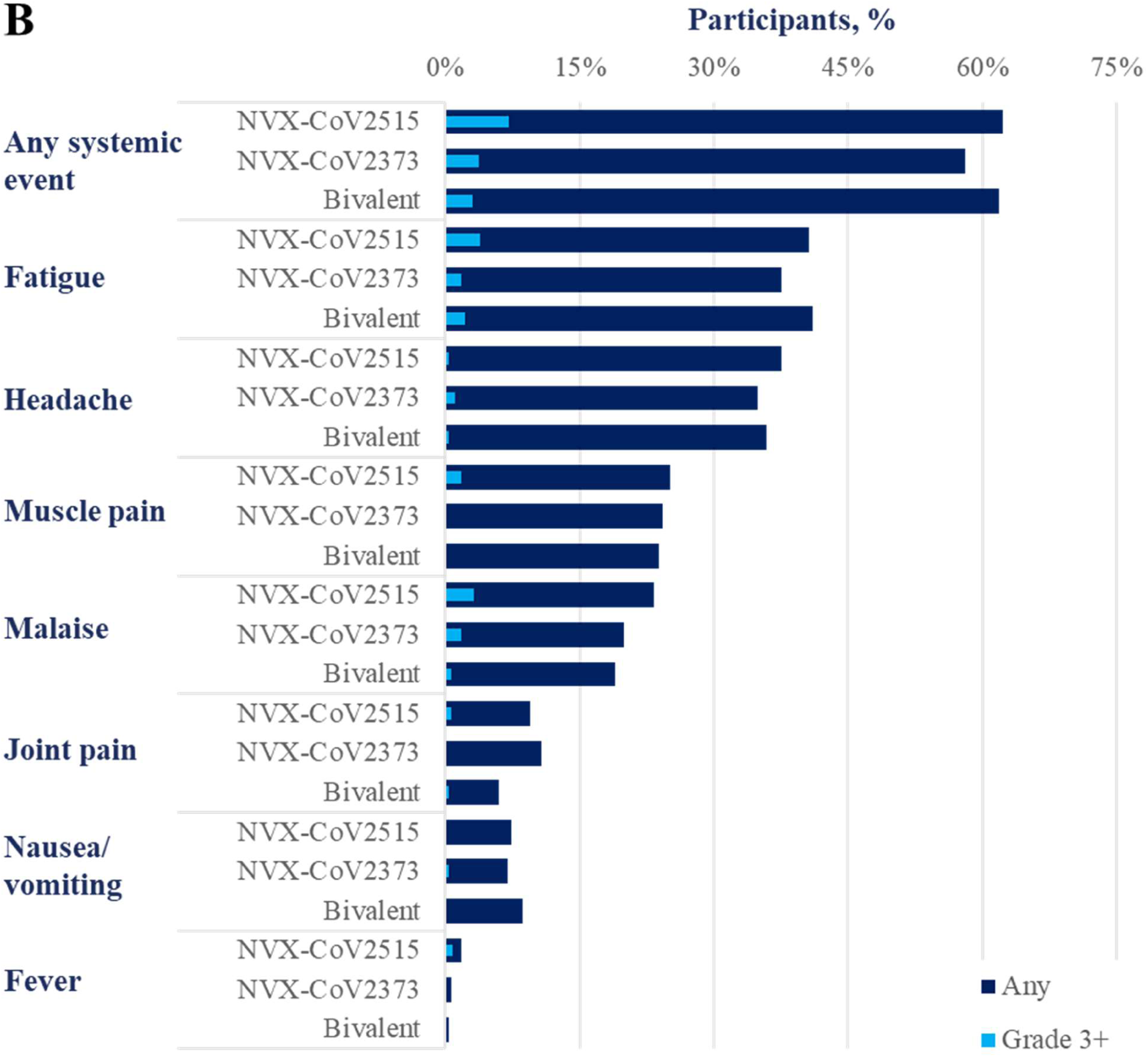
Solicited adverse events within 7 days of vaccination (Safety Analysis Set). (A) Frequencies of solicited local treatment-emergent adverse events. (B) Frequencies of solicited systemic treatment-emergent adverse events.

Through 28 days after vaccination, unsolicited TEAEs occurred in 34.3% (98/286), 38.0% (104/274), and 33.8% (91/269) of participants in the NVX-CoV2515, NVX-CoV2373, and the bivalent vaccine groups, respectively (**Table 4**). Unsolicited serious TEAEs occurred in 0.3% (1/286) and 0.4% (1/274) of participants in the NVX-CoV2515 and NVX-CoV2373, respectively, though neither of these events were reported as vaccine related. No unsolicited serious TEAEs occurred in the bivalent vaccine group. There were no treatment-related medically attended adverse events (MAAEs) or potential immune-mediated medical conditions (PIMMCs) in any vaccine group.

**Table 4.**
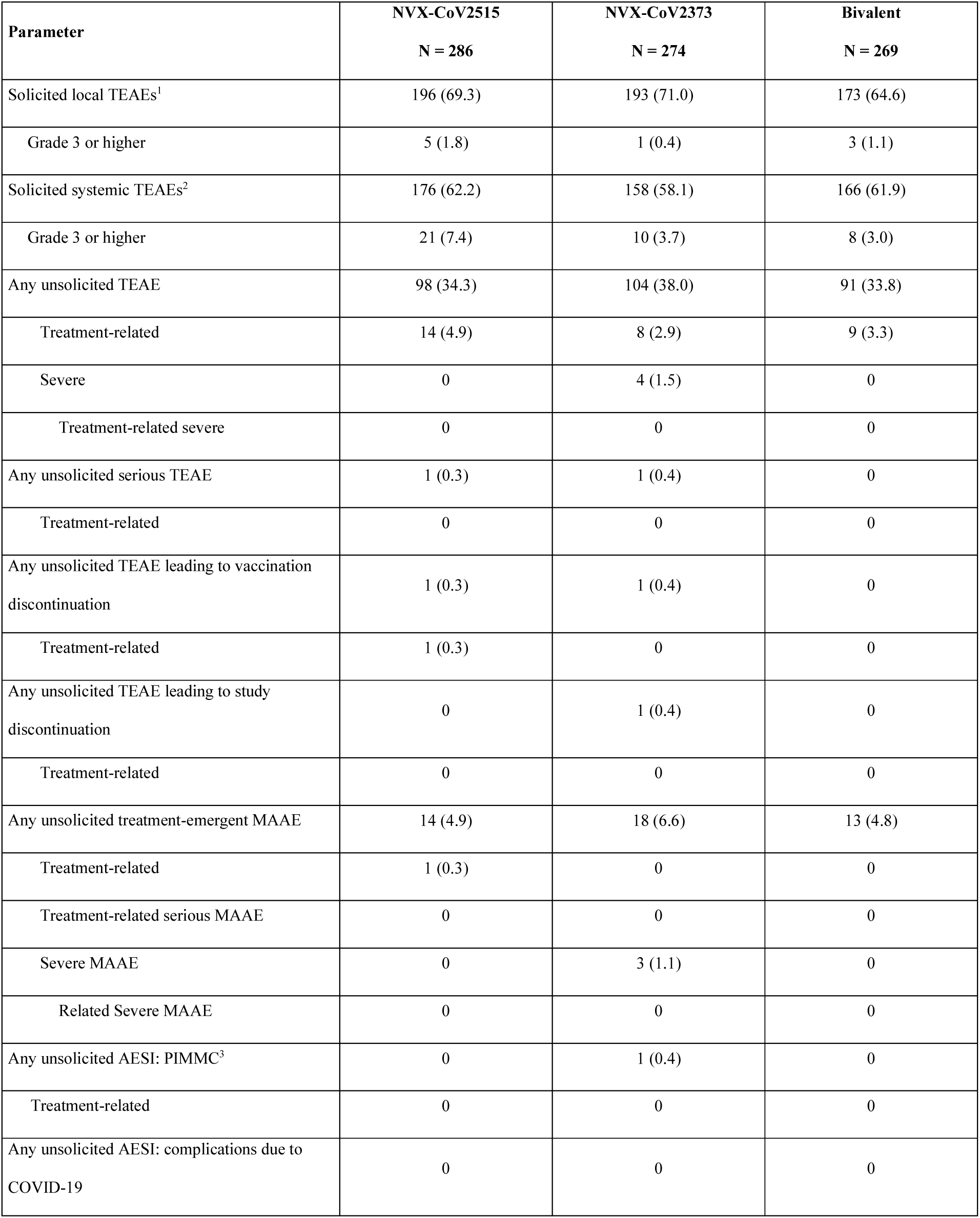
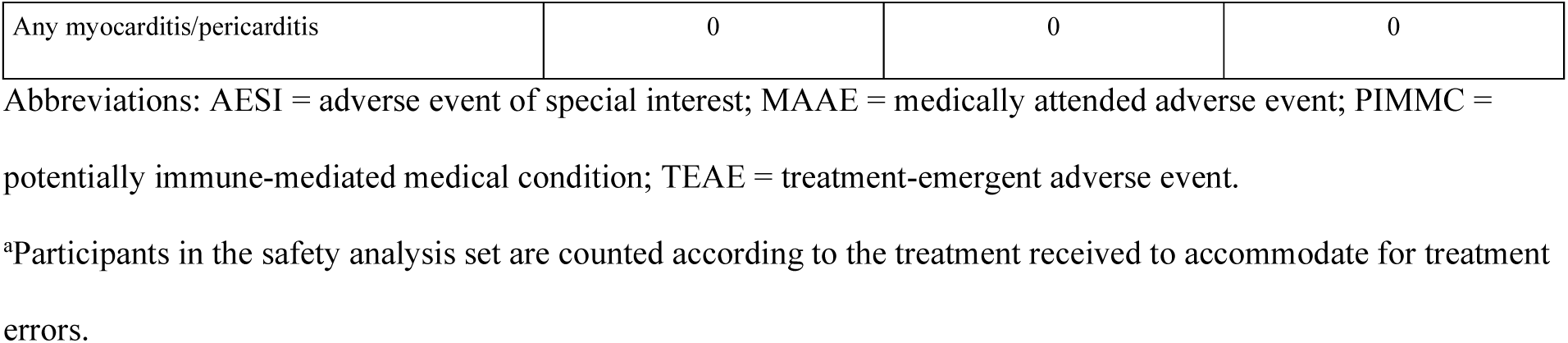
Overall summary of unsolicited treatment-emergent adverse events through 28 days after vaccination (Safety Analysis Set^a^)

| **Parameter** | **NVX-CoV2515**<br>**N = 286** | **NVX-CoV2373**<br>**N = 274** | **Bivalent**<br>**N = 269** |
| --- | --- | --- | --- |
| Solicited local TEAEs<sup>1</sup> | 196 (69.3) | 193 (71.0) | 173 (64.6) |
| Grade 3 or higher | 5 (1.8) | 1 (0.4) | 3 (1.1) |
| Solicited systemic TEAEs<sup>2</sup> | 176 (62.2) | 158 (58.1) | 166 (61.9) |
| Grade 3 or higher | 21 (7.4) | 10 (3.7) | 8 (3.0) |
| Any unsolicited TEAE | 98 (34.3) | 104 (38.0) | 91 (33.8) |
| Treatment-related | 14 (4.9) | 8 (2.9) | 9 (3.3) |
| Severe | 0 | 4 (1.5) | 0 |
| Treatment-related severe | 0 | 0 | 0 |
| Any unsolicited serious TEAE | 1 (0.3) | 1 (0.4) | 0 |
| Treatment-related | 0 | 0 | 0 |
| Any unsolicited TEAE leading to vaccination discontinuation | 1 (0.3) | 1 (0.4) | 0 |
| Treatment-related | 1 (0.3) | 0 | 0 |
| Any unsolicited TEAE leading to study discontinuation | 0 | 1 (0.4) | 0 |
| Treatment-related | 0 | 0 | 0 |
| Any unsolicited treatment-emergent MAAE | 14 (4.9) | 18 (6.6) | 13 (4.8) |
| Treatment-related | 1 (0.3) | 0 | 0 |
| Treatment-related serious MAAE | 0 | 0 | 0 |
| Severe MAAE | 0 | 3 (1.1) | 0 |
| Related Severe MAAE | 0 | 0 | 0 |
| Any unsolicited AESI: PIMMC<sup>3</sup> | 0 | 1 (0.4) | 0 |
| Treatment-related | 0 | 0 | 0 |
| Any unsolicited AESI: complications due to COVID-19 | 0 | 0 | 0 |
| Any myocarditis/pericarditis | 0 | 0 | 0 |

Abbreviations: AESI = adverse event of special interest; MAAE = medically attended adverse event; PIMMC = potentially immune-mediated medical condition; TEAE = treatment-emergent adverse event.
<sup>a</sup>Participants in the safety analysis set are counted according to the treatment received to accommodate for treatment errors.

## Discussion

In this report, we describe the first immunogenicity and safety data for Omicron-specific (NVX- CoV2515) and bivalent SARS-CoV-2 rS protein subunit vaccines. These results are from an interim analysis of an ongoing phase 3, randomized, observer-blinded, clinical trial in participants who previously received a regimen of 3 doses of prototype mRNA vaccine.

After participants received a single booster dose of either NVX-CoV2515, NVX-CoV2373, or the bivalent vaccine (NVX-CoV2515 + NVX-CoV2373), immunogenicity assessments— microneutralization (MN50), anti-S IgG (GMEU), and pseudovirus neutralization (ID50) assays— were analyzed on Days 14 and 28 post-vaccination.

The primary endpoint analysis demonstrated that the Omicron BA.1-specific vaccine, NVX- CoV2515, produced a superior neutralizing antibody response (MN50) against the Omicron BA.1 subvariant when compared with the prototype vaccine, NVX-CoV2373, and met the non-inferiority criterion for SRR versus NVX-CoV2373 at Day 14 following booster administration, thereby successfully achieving the study’s primary objective.

Although NVX-CoV2515 demonstrated superior MN50 titers against the Omicron BA.1 strain, when compared with NVX-CoV2373, this trend did not persist through all assay types. NVX- CoV2515, NVX-CoV2373, and the bivalent vaccine elicited similar MN50 and pseudovirus ID50 responses against the ancestral strain. Trends with the MN50 assays were maintained both in participants with no evidence of previous infection (PP1) and in participants with evidence of previous infection, as determined by anti-NP status at baseline (PP2). With regard to prevention of severe disease and hospitalization due to COVID-19, both neutralizing and non-neutralizing antibodies play a role in vaccine efficacy [17]. Thus, lower titers for neutralizing antibodies do not necessarily indicate lower vaccine efficacy. Overall, these assays revealed no consistent immunogenic differences between the prototype and the bivalent vaccine.

In a recent study, the immunogenicity of the mRNA-1273.214 bivalent vaccine (ancestral SARS-CoV-2 strain + Omicron BA.1) was investigated in individuals who previously received three doses of the prototype mRNA-1273 vaccine [11]. Compared with individuals who received the prototype mRNA.1273 vaccine as a fourth booster dose, those who received mRNA.1273.214 as a fourth dose exhibited higher binding antibody responses against Omicron BA.1 and Omicron BA.4/5 variants, resulting in the acknowledged superiority of mRNA.1273.214 to the mRNA- 1273 prototype vaccine. An additional study conducted with the same bivalent mRNA vaccine showed that both the Omicron-BA.1-monovalent mRNA-1273.529 and bivalent mRNA- 1273.214 vaccines elicited superior neutralizing antibody responses against Omicron BA.1 compared with the prototype mRNA-1273 vaccine [18].

After receiving a single dose of either NVX-CoV2515, NVX-CoV2373, or bivalent vaccine, participant sera from each of the 3 study groups achieved anti-Spike IgG antibody concentrations against the ancestral SARS-CoV-2 strain that were previously associated with vaccine efficacy levels of 88%-95% in pivotal phase 3 studies of the prototype vaccine [6,19]. Anti-Spike IgG antibody responses using the Omicron BA.1–specific assay were balanced across the 3 study groups, regardless of baseline status of confirmed prior infection, displaying similar benefits of NVX-CoV2515, the bivalent vaccine, and the prototype vaccine. The consistent anti-Spike IgG responses (agnostic of strain) suggest the development of broadly cross-reacting IgG antibodies following administration of SARS-CoV-2 rS protein subunit vaccines, either as the prototype, BA.1 variant, or bivalent vaccine.

Overall, the variant specific vaccine (NVX-CoV2515) induced a superior neutralizing antibody response against the Omicron BA.1 subvariant when compared with the prototype vaccine, NVX-CoV2373. The NVX-CoV2515 and bivalent SARS-CoV-2 rS protein subunit vaccines demonstrated similar immunogenicity 14 days post-vaccination, with no added benefits when compared with the prototype product (NVX-CoV2373) across several ancestral strain- and Omicron BA.1–specific immunoassays. Immunogenicity results at Day 28 were generally similar to those at Day 14.

The PP2 population assessed in this study most accurately represents a real-world population in which previous SARS-CoV-2 infection is not uncommon. In Australia as of November 2022, at least 66% of the population were estimated to had been previously infected by SARS-CoV-2 [20]. Anti-S IgG antibody responses against the Omicron BA.1 subvariant for the PP2 population were generally 1.5- to 2-fold greater than those for the PP1 population. These results align with studies that assessed immune responses in both participants with and without previous SARS-CoV-2 infection that showed that neutralization of subvariant SARS-CoV-2 strains was higher after a booster with bivalent mRNA vaccine than after a booster dose with prototype mRNA vaccine [10,11].

The incidence of solicited local and systemic reactogenicity reported in this study was consistent with previous studies of NVX-CoV2373, with pain/tenderness being the most common local solicited AE and fatigue being the most common solicited systemic AE [6,21]. Incidence rates for all local and systemic events were similar across all vaccine groups.

Incidences of unsolicited TEAEs and serious AEs were also unremarkable with respect to prior research on the prototype vaccine, and there were no reports of related MAAEs, PIMMCs, or SAEs. Collectively, these data were consistent with the safety profile of other variant-specific SARS-CoV-2 rS protein subunit vaccines, bivalent combination, or with use as a heterologous booster in combination with mRNA vaccines.

Our study was subject to certain limitations. As these results are from an ongoing phase 3 study conducted with a limited sample size, clinical efficacy of the booster dose was not evaluated.

Safety follow-up in this study, at present, is also limited to 28 days. Furthermore, it remains to be seen if the conclusions based on the prototype versus variant-specific vaccines in this study can be extrapolated to newer strains or newer vaccines. For example, in a recent study comparing the neutralization activity of a bivalent BA.4/5 BNT162b2 vaccine to the prototype BNT162b2 vaccine against newly emerged Omicron sublineages descended from BA.2 and BA.4/BA.5 in persons who previously received three doses of BNT162b2, the bivalent BA.4/5 vaccine was more immunogenic than the original BNT162b2 monovalent vaccine against circulating Omicron sublineages [10]. Additionally, newer Omicron subvariants such as BQ and XBB show marked evasion of vaccine-induced neutralization and evasion from monoclonal antibodies with known neutralization capability against the original Omicron variant [22]. Therefore, responses to the Omicron BA.5 variant after immunization with NVX-CoV2515, NVX-CoV2373, or bivalent vaccine, and the effect of a subsequent booster dose at 3 months, will be addressed in future work, given that BA.5 is more closely related phylogenetically to XBB and BQ than to the ancestral strain.

In conclusion, the variant-specific vaccine NVX-CoV2515 demonstrated superior neutralizing response against the matched Omicron BA.1 subvariant virus. The prototype and bivalent vaccines also induced robust immune responses to ancestral and Omicron subvariant strains of SARS-CoV-2 when administered as a fourth dose. Moreover, the safety profile of updated variant-specific SARS-CoV-2 rS protein subunit vaccines remained consistent with the prototype vaccine when administered as a heterologous booster dose following 3 vaccinations with mRNA vaccines.

## Supporting information

CONSORT Checklist

## Data Availability

Study information is available at https://clinicaltrials.gov/ct2/show/NCT05372588, and requests will be considered.

## Acknowledgments

We thank all the study participants who volunteered for this study. This study was funded by Novavax, Inc. Editorial support was provided by Kelly Cameron, PhD, and Amanda Cox, PhD, of Ashfield MedComms (New York, USA), an Inizio company, supported by Novavax, Inc.

## Contributors

RMM, KA, JSP, and NP were involved in the study design, data collection, and interpretation. SG performed the statistical analyses. All authors reviewed, commented on, and approved this manuscript prior to submission for publication. The authors were not precluded from accessing data in the study, and they accept responsibility to submit the manuscript for publication.

## References

1. Marks P. Coronavirus (COVID-19) Update: FDA Recommends Inclusion of Omicron BA.4/5 Component for COVID-19 Vaccine Booster Doses: U.S. Food & Drug Administration; 2022. Available from: https://www.fda.gov/news-events/press-announcements/coronavirus-covid-19-update-fda-recommends-inclusion-omicron-ba45-component-covid-19-vaccine-booster

2. Centers for Disease Control and Prevention. SARS-CoV-2 variant classifications and definitions [updated 2023 March 20; cited 2023 May 5]. Available from: https://www.cdc.gov/coronavirus/2019-ncov/variants/variant-classifications.html#anchor_1632154493691

3. Polack FP, Thomas SJ, Kitchin N, et al. Safety and Efficacy of the BNT162b2 mRNA Covid-19 Vaccine. N Engl J Med. 2020 Dec 31;383(27):2603–2615.

4. Baden LR, El Sahly HM, Essink B, et al. Efficacy and Safety of the mRNA-1273 SARS-CoV-2 Vaccine. N Engl J Med. 2021 Feb 4;384(5):403–416.

5. Heath PT, Galiza EP, Baxter DN, et al. Safety and Efficacy of NVX-CoV2373 Covid-19 Vaccine. N Engl J Med. 2021 Sep 23;385(13):1172-1183.

6. Dunkle LM, Kotloff KL, Gay CL, et al. Efficacy and Safety of NVX-CoV2373 in Adults in the United States and Mexico. N Engl J Med. 2022 Feb 10;386(6):531–543.

7. Tartof SY, Slezak JM, Puzniak L, et al. BNT162b2 vaccine effectiveness against SARS-CoV-2 omicron BA.4 and BA.5. Lancet Infect Dis. 2022 Dec;22(12):1663–1665.

8. Kurhade C, Zou J, Xia H, et al. Neutralization of Omicron BA.1, BA.2, and BA.3 SARS-CoV-2 by 3 doses of BNT162b2 vaccine. Nat Commun. 2022 Jun 23;13(1):3602.

9. Tseng HF, Ackerson BK, Bruxvoort KJ, et al. Effectiveness of mRNA-1273 vaccination against SARS-CoV-2 omicron subvariants BA.1, BA.2, BA.2.12.1, BA.4, and BA.5. Nat Commun. 2023 Jan 12;14(1):189.

10. Zou J, Kurhade C, Patel S, et al. Neutralization of BA.4-BA.5, BA.4.6, BA.2.75.2, BQ.1.1, and XBB.1 with Bivalent Vaccine. N Engl J Med. 2023 Mar 2;388(9):854–857.

11. Chalkias S, Harper C, Vrbicky K, et al. A Bivalent Omicron-Containing Booster Vaccine against Covid-19. N Engl J Med. 2022 Oct 6;387(14):1279–1291.

12. US Food and Drug Administration (FDA). FDA Briefing Document: Vaccines and Related Biological Products Advisory Committee Meeting. Selection of Strain(s) to be Included in the Periodic Updated COVID-19 Vaccines for the 2023-2024 Vaccination Campaign. 2023 June 15.

13. World Health Organization (WHO). Statement on the antigen composition of COVID-19 vaccines 2023 May 18 [cited 2023 June 27]. Available from: https://www.who.int/news/item/18-05-2023-statement-on-the-antigen-composition-of-covid-19-vaccines

14. European Medicines Agency (EMA). EMA and ECDC statement on updating COVID-19 Vaccines to target new SARS-CoV-2 virus variants 2023 June 6 [cited 2023 June 27]. Available from: https://www.ema.europa.eu/en/news/ema-ecdc-statement-updating-covid-19-vaccines-target-new-sars-cov-2-virus-variants#:~:text=In%20line%20with%20the%20outcome,other%20parts%20of%20the%20world.

15. Madhi SA, Moodley D, Hanley S, et al. Immunogenicity and safety of a SARS-CoV-2 recombinant spike protein nanoparticle vaccine in people living with and without HIV-1 infection: a randomised, controlled, phase 2A/2B trial. Lancet HIV. 2022 May;9(5):e309–e322.

16. Huang Y, Borisov O, Kee JJ, et al. Calibration of two validated SARS-CoV-2 pseudovirus neutralization assays for COVID-19 vaccine evaluation. Sci Rep. 2021 Dec 14;11(1):23921.

17. Bates TA, Lu P, Kang YJ, et al. BNT162b2-induced neutralizing and non-neutralizing antibody functions against SARS-CoV-2 diminish with age. Cell Rep. 2022 Oct 25;41(4):111544.

18. Lee IT, Cosgrove CA, Moore P, et al. A Randomized Trial Comparing Omicron-Containing Boosters with the Original Covid-19 Vaccine mRNA-1273. medRxiv. 2023.

19. Fong Y, Huang Y, Benkeser D, et al. Immune correlates analysis of the PREVENT-19 COVID-19 vaccine efficacy clinical trial. Nat Commun. 2023 Jan 19;14(1):331.

20. Australian COVID-19 Serosurveillance Network. Seroprevalence of SARS-CoV-2-specific antibodies among Australian blood donors: Round 3 update. [updated 03 November 2022; cited 2023 14 April]. Available from: https://kirby.unsw.edu.au/sites/default/files/COVID19-Blood-Donor-Report-Round3-Aug-Sep-2022.pdf

21. Mallory RM, Formica N, Pfeiffer S, et al. Safety and immunogenicity following a homologous booster dose of a SARS-CoV-2 recombinant spike protein vaccine (NVX-CoV2373): a secondary analysis of a randomised, placebo-controlled, phase 2 trial. Lancet Infect Dis. 2022 Nov;22(11):1565–1576.

22. Wang Q, Iketani S, Li Z, et al. Alarming antibody evasion properties of rising SARS- CoV-2 BQ and XBB subvariants. Cell. 2023 Jan 19;186(2):279–286 e8.

