## Supplemental Materials for "Immunogenicity and safety of heterologous Omicron BA.1 and bivalent SARS-CoV-2 recombinant spike protein booster vaccines: a phase 3, randomized, clinical trial"

### Supplemental Tables and Figures

#### Supplemental Table 1. Demographics and baseline disease characteristics (PP1 and PP2 Analysis Sets)

| Parameters | PP1 Analysis Set | | | PP2 Analysis Set | | |
|---|---|---|---|---|---|---|
| | NVX-CoV2515 N = 126 | NVX-CoV2373 N = 119 | Bivalent (NVX-CoV2373 + NVX-CoV2515) N = 118 | NVX-CoV2515 N = 258 | NVX-CoV2373 N = 251 | Bivalent (NVX-CoV2373 + NVX-CoV2515) N = 240 |
| **Age (years)** | | | | | | |
| Mean (SD) | 42.0 (11.73) | 42.1 (11.10) | 42.0 (12.18) | 40.5 (12.4) | 40.0 (11.5) | 39.7 (12.5) |
| Median | 44.5 | 43.0 | 42.5 | 42.0 | 41.0 | 41.0 |
| Min–max | 20–64 | 18–63 | 18–63 | 18–64 | 18–64 | 18–64 |
| **Sex, n (%)** | | | | | | |
| Male | 47 (37.3) | 52 (43.7) | 52 (44.1) | 119 (46.1) | 122 (48.6) | 108 (45.0) |
| Female | 79 (62.7) | 67 (56.3) | 66 (55.9) | 139 (53.9) | 129 (51.4) | 132 (55.0) |
| **Race, n (%)** | | | | | | |
| White | 108 (85.7) | 98 (82.4) | 100 (84.7) | 208 (80.6) | 195 (77.7) | 195 (81.3) |
| Black or African American | 0 | 2 (1.7) | 0 | 0 | 2 (0.8) | 0 |
| Aboriginal Australian | 1 (0.8) | 0 | 0 | 2 (0.8) | 0 | 0 |
| Native Hawaiian or Other Pacific Islander | 0 | 0 | 1 (0.8) | 1 (0.4) | 0 | 1 (0.4) |

**Supplemental Table 1. Demographics and baseline disease characteristics (PP1 and PP2 Analysis Sets)**

| Parameters | PP1 Analysis Set | | | PP2 Analysis Set | | |
|---|---|---|---|---|---|---|
| | NVX-CoV2515 N = 126 | NVX-CoV2373 N = 119 | Bivalent (NVX-CoV2373 + NVX-CoV2515) N = 118 | NVX-CoV2515 N = 258 | NVX-CoV2373 N = 251 | Bivalent (NVX-CoV2373 + NVX-CoV2515) N = 240 |
| Asian | 14 (11.1) | 15 (12.6) | 17 (14.4) | 35 (13.6) | 43 (17.1) | 37 (15.4) |
| Mixed Origin | 2 (1.6) | 3 (2.5) | 0 | 5 (1.9) | 3 (1.2) | 1 (0.4) |
| Other | 1 (0.8) | 1 (0.8) | 0 | 7 (2.7) | 8 (3.2) | 6 (2.5) |
| Not Reported | 0 | 0 | 0 | 0 | 0 | 0 |
| **Ethnicity, n (%)** | | | | | | |
| Australian | 116 (92.1) | 105 (88.2) | 108 (91.5) | 229 (88.8) | 217 (86.5) | 209 (87.1) |
| Aboriginal/Torres Strait Islanders | 1 (0.8) | 2 (1.7) | 0 | 4 (1.6) | 2 (0.8) | 0 |
| Hispanic or Latino | 1 (0.8) | 1 (0.8) | 1 (0.8) | 5 (1.9) | 7 (2.8) | 6 (2.5) |
| Not reported | 5 (4.0) | 5 (4.2) | 5 (4.2) | 9 (3.5) | 14 (5.6) | 14 (5.8) |
| Unknown | 3 (2.4) | 6 (5.0) | 3 (2.5) | 9 (3.5) | 10 (4.0) | 9 (3.8) |
| Missing | 0 | 0 | 1 (0.8) | 2 (0.8) | 1 (0.4) | 2 (0.8) |
| **BMI (kg/m$^2$)** | | | | | | |
| Mean (SD) | 28.89 (6.900) | 28.06 (5.191) | 27.40 (5.841) | 28.2 (6.5) | 27.9 (5.2) | 27.4 (5.7) |
| Median | 27.40 | 27.50 | 26.20 | 27.20 | 27.50 | 26.30 |

**Supplemental Table 1. Demographics and baseline disease characteristics (PP1 and PP2 Analysis Sets)**

| Parameters | PP1 Analysis Set | | | PP2 Analysis Set | | |
|---|---|---|---|---|---|---|
| | NVX-CoV2515 N = 126 | NVX-CoV2373 N = 119 | Bivalent (NVX-CoV2373 + NVX-CoV2515) N = 118 | NVX-CoV2515 N = 258 | NVX-CoV2373 N = 251 | Bivalent (NVX-CoV2373 + NVX-CoV2515) N = 240 |
| Min–max | 18.4–54.6 | 17.9–47.2 | 18.2–50.1 | 18.1–55.8 | 17.4–47.2 | 17.9–50.1 |
| **BMI (kg/m$^2$) category, n (%)** | | | | | | |
| Underweight (<18.0) | 0 | 1 (0.8) | 0 | 0 | 3 (1.2) | 1 (0.4) |
| Normal (18.0–24.9) | 41 (32.5) | 32 (26.9) | 46 (39.0) | 93 (36.0) | 69 (27.5) | 94 (39.2) |
| Overweight (25.0–29.9) | 39 (31.0) | 49 (41.2) | 46 (39.0) | 80 (31.0) | 100 (39.8) | 82 (34.2) |
| Obese (≥ 30.0) | 46 (36.5) | 37 (31.1) | 26 (22.0) | 84 (32.6) | 76 (30.3) | 61 (25.4) |
| Missing | 0 | 0 | 0 | 1 (0.4) | 3 (1.2) | 2 (0.8) |
| **Regimen of previous COVID-19 vaccine, n (%)** | | | | | | |
| Moderna | 0 | 0 | 3 (2.5) | 0 | 2 (0.8) | 5 (2.1) |
| Pfizer-BioNTech | 95 (75.4) | 90 (75.6) | 89 (75.4) | 199 (77.1) | 194 (77.3) | 175 (72.9) |
| Mixed | 31 (24.6) | 29 (24.4) | 26 (22.0) | 59 (22.9) | 55 (21.9) | 60 (25.0) |
| Moderna-Moderna-Pfizer | 0 | 1 (0.8) | 0 | 1 (0.4) | 1 (0.4) | 0 |
| Moderna-Pfizer-Pfizer | 2 (1.6) | 0 | 0 | 2 (0.8) | 0 | 1 (0.4) |
| Moderna-Pfizer-Moderna | 0 | 0 | 0 | 0 | 0 | 0 |

**Supplemental Table 1. Demographics and baseline disease characteristics (PP1 and PP2 Analysis Sets)**

| Parameters | PP1 Analysis Set | | | PP2 Analysis Set | | |
|---|---|---|---|---|---|---|
| | NVX-CoV2515 N = 126 | NVX-CoV2373 N = 119 | Bivalent (NVX-CoV2373 + NVX-CoV2515) N = 118 | NVX-CoV2515 N = 258 | NVX-CoV2373 N = 251 | Bivalent (NVX-CoV2373 + NVX-CoV2515) N = 240 |
| Pfizer-Pfizer-Moderna | 29 (23.0) | 28 (23.5) | 26 (22.0) | 56 (21.7) | 53 (21.1) | 59 (24.6) |
| Pfizer-Moderna-Moderna | 0 | 0 | 0 | 0 | 1 (0.4) | 0 |
| Pfizer-Moderna-Pfizer | 0 | 0 | 0 | 0 | 0 | 0 |
| **Previous COVID-19, n (%)** | | | | | | |
| Yes | 0 | 1 (0.8) | 1 (0.8) | 15 (5.8) | 16 (6.4) | 15 (6.3) |
| No | 126 (100) | 118 (99.2) | 117 (99.2) | 243 (94.2) | 235 (93.6) | 225 (93.8) |
| **Qualitative anti-N, n (%)** | | | | | | |
| Positive | 0 | 0 | 0 | 132 (51.2) | 132 (52.6) | 122 (50.8) |
| Negative | 126 (100) | 119 (100) | 118 (100) | 126 (48.8) | 119 (47.4) | 118 (49.2) |
| **PCR, n (%)** | | | | | | |
| Positive | 0 | 0 | 0 | 0 | 0 | 0 |
| Negative | 126 (100) | 119 (100) | 118 (100) | 258 (100) | 251 (100) | 240 (100) |
| **Anti-N / PCR, n (%)[1]** | | | | | | |
| Positive | 0 | 0 | 0 | 132 (51.2) | 132 (52.6) | 122 (50.8) |

**Supplemental Table 1. Demographics and baseline disease characteristics (PP1 and PP2 Analysis Sets)**

| Parameters | PP1 Analysis Set | | | PP2 Analysis Set | | |
|---|---|---|---|---|---|---|
| | NVX-CoV2515 N = 126 | NVX-CoV2373 N = 119 | Bivalent (NVX-CoV2373 + NVX-CoV2515) N = 118 | NVX-CoV2515 N = 258 | NVX-CoV2373 N = 251 | Bivalent (NVX-CoV2373 + NVX-CoV2515) N = 240 |
| Negative | 126 (100) | 119 (100) | 118 (100) | 126 (48.8) | 119 (47.4) | 118 (49.2) |
| **Time between last previous COVID-19 vaccine and booster dose of study vaccine (days)** | | | | | | |
| Mean (SD) | 181.6 (39.09) | 181.5 (32.02) | 181.7 (32.24) | 177.7 (39.4) | 181.8 (35.9) | 178.8 (36.7) |
| Median | 178.0 | 181.0 | 182.5 | 177.0 | 182.0 | 180.0 |
| Min–max | 105–440 | 91–267 | 110–306 | 84–440 | 91–329 | 91–313 |
| **Interval between last previous COVID-19 vaccine and booster dose of study vaccine, n (%)** | | | | | | |
| <90 days | 0 | 0 | 0 | 1 (0.4) | 0 | 0 |
| 90–120 days | 4 (3.2) | 4 (3.4) | 4 (3.4) | 15 (5.8) | 15 (6.0) | 17 (7.1) |
| >120–150 days | 17 (13.5) | 16 (13.4) | 15 (12.7) | 38 (14.7) | 31 (12.4) | 33 (13.8) |
| >150–180 days | 49 (38.9) | 38 (31.9) | 36 (30.5) | 87 (33.7) | 73 (29.1) | 72 (30.0) |
| >180–210 days | 38 (30.2) | 43 (36.1) | 46 (39.0) | 80 (31.0) | 89 (35.5) | 82 (34.2) |
| >210–240 days | 10 (7.9) | 13 (10.9) | 12 (10.2) | 23 (8.9) | 31 (12.4) | 24 (10.0) |
| >240–270 days | 5 (4.0) | 5 (4.2) | 4 (3.4) | 8 (3.1) | 9 (3.6) | 9 (3.8) |
| >270–300 days | 2 (1.6) | 0 | 0 | 4 (1.6) | 1 (0.4) | 1 (0.4) |

**Supplemental Table 1. Demographics and baseline disease characteristics (PP1 and PP2 Analysis Sets)**

| Parameters | PP1 Analysis Set | | | PP2 Analysis Set | | |
|---|---|---|---|---|---|---|
| | NVX-CoV2515 N = 126 | NVX-CoV2373 N = 119 | Bivalent (NVX-CoV2373 + NVX-CoV2515) N = 118 | NVX-CoV2515 N = 258 | NVX-CoV2373 N = 251 | Bivalent (NVX-CoV2373 + NVX-CoV2515) N = 240 |
| >300–330 days | 0 | 0 | 1 (0.8) | 1 (0.4) | 2 (0.8) | 2 (0.8) |
| >330–360 days | 0 | 0 | 0 | 0 | 0 | 0 |
| >360 days | 1 (0.8) | 0 | 0 | 1 (0.4) | 0 | 0 |

Abbreviations: anti-N = anti-nucleocapsid; BMI = body mass index; COVID-19 = coronavirus disease 2019; max = maximum; min = minimum; NVX-CoV2515 = 5 µg SARS-CoV-2 rS with 50 µg Matrix-M™ adjuvant; NVX-CoV2373 = 5 µg SARS-CoV-2 rS with 50 µg Matrix-M adjuvant; NVX-CoV2373 + NVX-CoV2515 = 5 µg SARS-CoV-2 rS with 50 µg Matrix-M adjuvant (total); PCR = polymerase chain reaction; SARS-CoV-2 = severe acute respiratory syndrome coronavirus 2; SARS-CoV-2 rS = severe acute respiratory syndrome coronavirus 2 recombinant spike protein nanoparticle vaccine; SD = standard deviation.

Note: Age was calculated at the time of informed consent.

Note: n for continuous parameters represents the number of participants with non-missing values for that parameter.

Note: BMI was calculated as weight (kg) divided by squared height (m). Percentages were based on the respective Per Protocol Analysis Set within each treatment and overall.

1. Participants with either anti-N or PCR are reported.

**Supplemental Figure 1. CONSORT diagram**

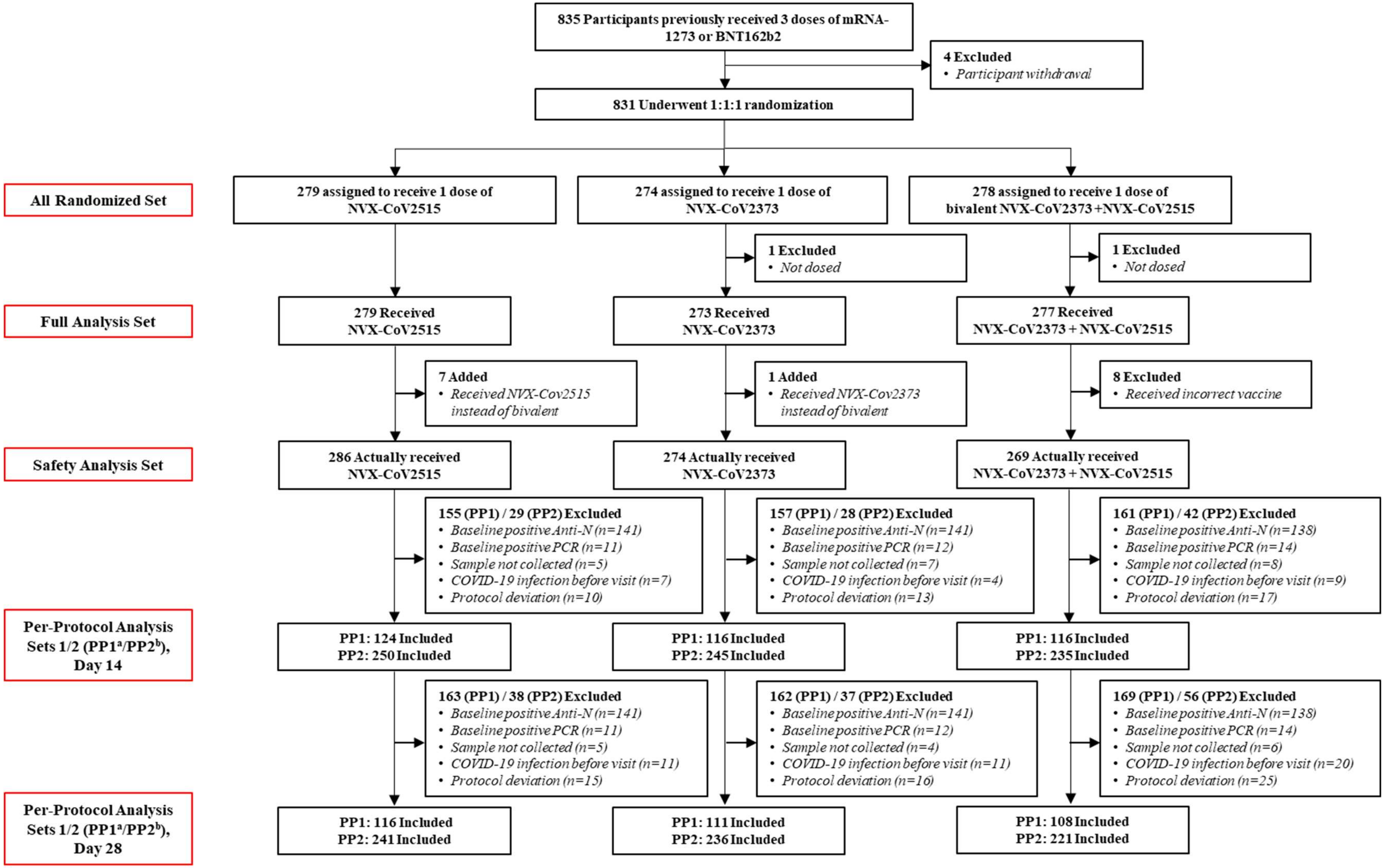


[a]PP1 includes all participants who received the full prescribed regimen of the study vaccine, had serology results for baseline and the time point analyzed, were

negative at baseline for SARS-CoV-2, and had no major protocol violations or an event (e.g., COVID-19 infection) that was considered clinically relevant to impact immunogenicity response. [b]PP2 population defined exactly as the PP1 population except that participants positive at baseline for SARS-CoV-2 were included.
